# Diffusion Kurtosis Imaging of neonatal Spinal Cord in clinical routine

**DOI:** 10.1101/2021.03.12.21253413

**Authors:** Rosella Trò, Monica Roascio, Domenico Tortora, Mariasavina Severino, Andrea Rossi, Julien Cohen-Adad, Marco Massimo Fato, Gabriele Arnulfo

**Affiliations:** Department of Informatics, Bioengineering, Robotics and System Engineering (DIBRIS), University of Genoa, Genoa, Italy; Neuroradiology Unit, Istituto Giannina Gaslini, Genoa, Italy; Department of Health Sciences (DISSAL), University of Genoa, Genoa, Italy; NeuroPoly Lab, Institute of Biomedical Engineering, Polytechnique Montreal, Montreal, QC, Canada; Functional Neuroimaging Unit, CRIUGM, Université de Montréal, Montreal, QC, Canada; Mila - Quebec AI Institute, Montreal, QC, Canada; Neuroscience Center, Helsinki Institute of Life Science, University of Helsinki, Helsinki, Finland

**Keywords:** Spinal Cord, DTI, DKI, image processing pipeline, neonatal imaging, punctate white matter lesions

## Abstract

Diffusion Kurtosis Imaging (DKI) has undisputed advantages over more classical diffusion Magnetic Resonance Imaging (dMRI), as witnessed by a fast-increasing number of clinical applications and software packages widely adopted in brain imaging. However, in the neonatal setting, DKI is still largely underutilized in Spinal Cord (SC) imaging because of its inherently demanding technological requirements.

Due to its extreme sensitivity to non-gaussian diffusion, DKI proves particularly suitable for detecting complex, subtle, fast microstructural changes occurring in this area at this early and critical stage of development, and not identifiable with only DTI. Given the multiplicity of congenital anomalies of the spinal canal, their crucial effect on later developmental outcome, and the close interconnection between SC region and the above brain, managing to apply such a method to neonatal cohort becomes of utmost importance.

This study will (i) review the current methodological challenges associated with application of advanced dMRI methods like DKI in early infancy, (ii) illustrate the first semi-automated pipeline built on Spinal Cord Toolbox (SCT) for handling with DKI data of neonatal SC, from acquisition setting to estimation of diffusion measures, through accurate adjustment of processing algorithms customized for adult SC, and (iii) present preliminary results of its application in a pilot clinical case study.

Results of this application agree with findings achieved in a corresponding adult survey, thus confirming validity of adopted pipeline and diagnostic value of DKI in pediatrics.

## 1. Introduction

In recent years, an increasing number of works in the field of neuroimaging are stressing the importance to move beyond simplistic assumptions of Diffusion Tensor Imaging (DTI) model (1) towards more advanced diffusion MRI (dMRI) methods, among which DKI (2) is one of the most promising (3–5).

Within existing non-standard techniques, DKI has indeed turned out to be especially suitable for imaging of SC, a structure where the assumption of Gaussian diffusion fails (6). Indeed, the presence of Gray Matter (GM) in the central portion contains a significant amount of cell membranes and organelles that limits diffusion to fewer directions. Taking into account pathological processes not following a Gaussian distribution, DKI provides a better understanding of the underlying micromolecular environment. In fact, it exhibits increased sensitivity in microstructural assessment of both White Matter (WM) and GM (7). Hence this susceptibility translates into an increased amount of diagnostic information, beyond that obtained with routine diffusion metrics.

Reduced field-of-view mitigates susceptibility artifacts and cardiac/respiratory gating allows to overcome most of the methodological challenges inherent to adult SC imaging (8). Thanks to these strategies, DKI by now represents a promising tool for studying a plethora of spine disorders with minor modifications to protocol parameters in use for brain imaging (9–14).

The scenario becomes definitely more complicated when attempting to translate this imaging technique to the pediatric clinical setting (15,16). Typical issues inherent to SC district include small cross-sectional area requiring high spatial resolution, interface between regions with different magnetic properties, Partial Volume Effect (PVE) of pulsating Cerebro-Spinal Fluid (CSF) with each heartbeat, and bulk physiologic motion due to proximity of heart and pulmonary parenchyma. This scenario is further complicated here from a multiplicity of factors related to the age range under analysis. Children in general have smaller anatomical structures - which in turn might result in higher risk of radiofrequency heating effects - and move more frequently (e.g. tongue sucking motion). On the other hand, artifact-reducing techniques (i.e. cardiac gating, respiratory compensation, and suppression sequences) (17) are often unfeasible since time-consuming, and sedation is typically not desirable. All aforementioned issues result in artifact-laden, low-signal images, which are often suboptimal for diagnostic evaluation.

Adopted solutions for improving image resolution and reducing artifacts comprise induction of natural sleep (by feeding the patient immediately before MR examination), the use of a vacuum fixation pillow to wrap the patient, and use of special ear muffs to protect from noise. However, the main requirement when handling with pediatric dMRI data is the choice of a proper acquisition protocol tailored for pediatric imaging, made up of low angular resolution, low b-values and few gradient directions, likewise in pediatric brain (18) in order to minimize scan time. Nevertheless, this forced time minimization clashes with specific requirements of advanced diffusion methods in terms of acquisition sequences.

Indeed, contrary to DTI, DKI and higher-order diffusion models requires multi-shell high angular resolution diffusion imaging (HARDI) sequences (19), typically involving at least three non-zero b-values distributed on hundreds of gradient directions, grouped in shells. This implies longer acquisition time, straining the feasibility of advanced dMRI methods in pediatrics. Resorting to optimized acquisition sequences (20), often combined with state-of-the-art techniques such as Parallel Imaging (21) and Multi-Band, can significantly increase acquisition speed and reduce artifacts. However, these advanced technologies are not always available in a general hospital due to high costs and technical limitations.

If extension of DTI to the pediatric SC has shown promising results in a wide range of clinical conditions, as evidenced by the increasing number of works on the topic (22–31), what immediately stands out while reviewing literature on pediatric SC is the absence of studies concerning DKI and particularly applied to the neonatal period (0-1 month).

To the best of our knowledge, the only published work on pediatric DKI (32) is limited to grown-up children (6-16 years), whose larger anatomical structures and reduced source of movements enable better image quality and longer scan times. Indeed, in newborns, SC dimensions themselves - 24 cm average length and 4.4 mm diameter, which can further decrease in case of malformations (33)-are enough to conceive amplification of aforementioned technical issues and thus justify the lack of research towards this direction.

However, the ability of DKI to offer additional and complementary information to DTI may bring a significant contribution in investigating such decisive and delicate stage of development, especially if we consider the wide range of developmental anomalies of the spinal canal affecting infants at birth (34–37).

It is on this premise that we conceived our work, whose aim is to show the feasibility of applying DKI to neonatal SC within clinical routine with all the issues that this entails.

We thus introduce here the first complete pipeline specifically adapted to neonatal imaging acquired for diagnostic purposes. Applicability and clinical validity of proposed method has been evaluated analyzing a specific clinical case-study concerning a condition common to preterm birth, in collaboration with Neuroradiology Unit of Giannina Gaslini Children’s Hospital of Genova.

Specifically, we assessed effects of WM brain lesions typical of Periventricular White Matter Injury (PWMI) on below cervical SC tracts by comparing diffusion measures between pathological patients and healthy controls. Our findings, though preliminary, confirm ability of DKI model in capturing subtle pathological alterations.

Since there are currently neither available protocols nor standardized methodological pipelines for performing DKI in the infant SC, this methodological outline may at least serve as a proof-of-concept, stressing the need for infant-specific data acquisition and processing guidelines in order to translate DKI of neonatal SC into routine clinical practice.

## 2. Materials and Methods

### 2.1 Subjects

Infants whose data have been used to disclose each step of the pipeline have been enrolled since August 2019 and scanned with 3.0 T MR scanner using a 32-channel head array coil (Ingenia Cx, Philips, Best, the Netherlands) at the Neuroradiology Unit of Giannina Gaslini Children’s Hospital of Genova. Conventional MRI and DKI were performed in 17 pre-term infants (28.1 to 36.7 weeks Gestational Age (GA); scanned at Term-Equivalent Age (TEA)). Details about subjects’ demographics are reported in Table 1.

**Table 1:**
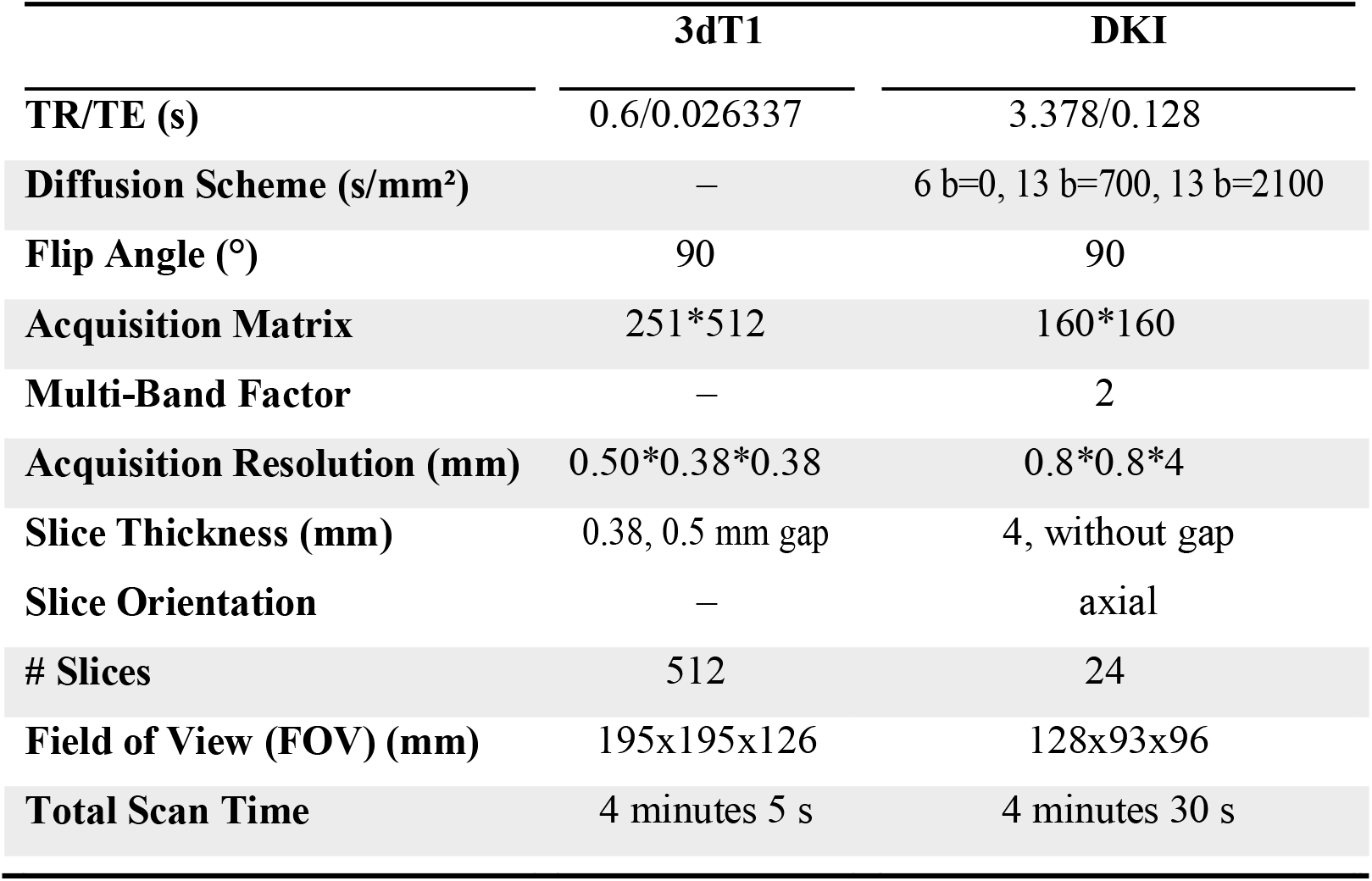
Demographics features of infants.

This single-center study was carried out in accordance with the recommendations of “Comitato Etico Regione Liguria, Genova, Italy” with written informed parental consent obtained for each infant prior to examination in accordance with the Declaration of Helsinki. Subjects were spontaneously breathing during examination; free-flowing oxygen was administered for all the duration of MRI session if necessary. Throughout the course of the examination, newborns were subjected to constant monitoring of the oxygen saturation and heart rate, by pulse oximeter and three-electrode electrocardiographic monitoring, respectively.

Exclusion criteria included obvious motion artifacts, oblique positioning, an incomplete imaging process or a low SNR. We opted for focusing exclusively on upper SC portion, by only acquiring the cervical district (C1-C7), affected by a significantly lower degree of physiologic motion than the dorsal and lumbar portions (38).

In consensus with a board-certified pediatric neuroradiologist, we performed QC for each of the pipeline’s steps.

### 2.2. Full pipeline description

Our pipeline integrates MRtrix3 (v.3.0.1) (39) for setting of dMRI acquisition sequence, Spinal Cord Toolbox (SCT, v. 5.3.0, https://github.com/neuropoly/spinalcordtoolbox) (40) for all processing steps specific to the SC, and Diffusion Imaging in Python (Dipy, v.1.4.0) (41) for denoising as well as computation of diffusion metrics.

Output of key processes, such as motion correction, segmentation and registration with atlas, can be checked through an interactive SCT Quality Control (QC) module, which automatically generates reports consisting in HTML files, containing a table of entries and allowing to show, for each entry, animated images (background with overlay on and off) for data quality validation.

In our methodological pipeline we have opted for mainly relying on SCT, being currently the only existing fully-comprehensive, free and open-source software dedicated to the processing and analysis of multi-parametric MRI of the spinal cord (42–53) successfully employed in a plethora of clinical applications concerning adult SC (54–63).

An overview of our image processing pipeline highlighting key features is shown in Figure 1. Since SCT algorithms are validated in adult imaging, we specifically customized each processing step to our neonatal scans. Our pipeline thus represents, to the best of our knowledge, the first semi-automated ad-hoc procedure for imaging of neonatal spine. A fully automatic workflow is not feasible here: acquisition time constraints, available scanner features, and subsequent image quality require inevitable although minimal and highly reproducible manual interventions.

**Figure 1.**
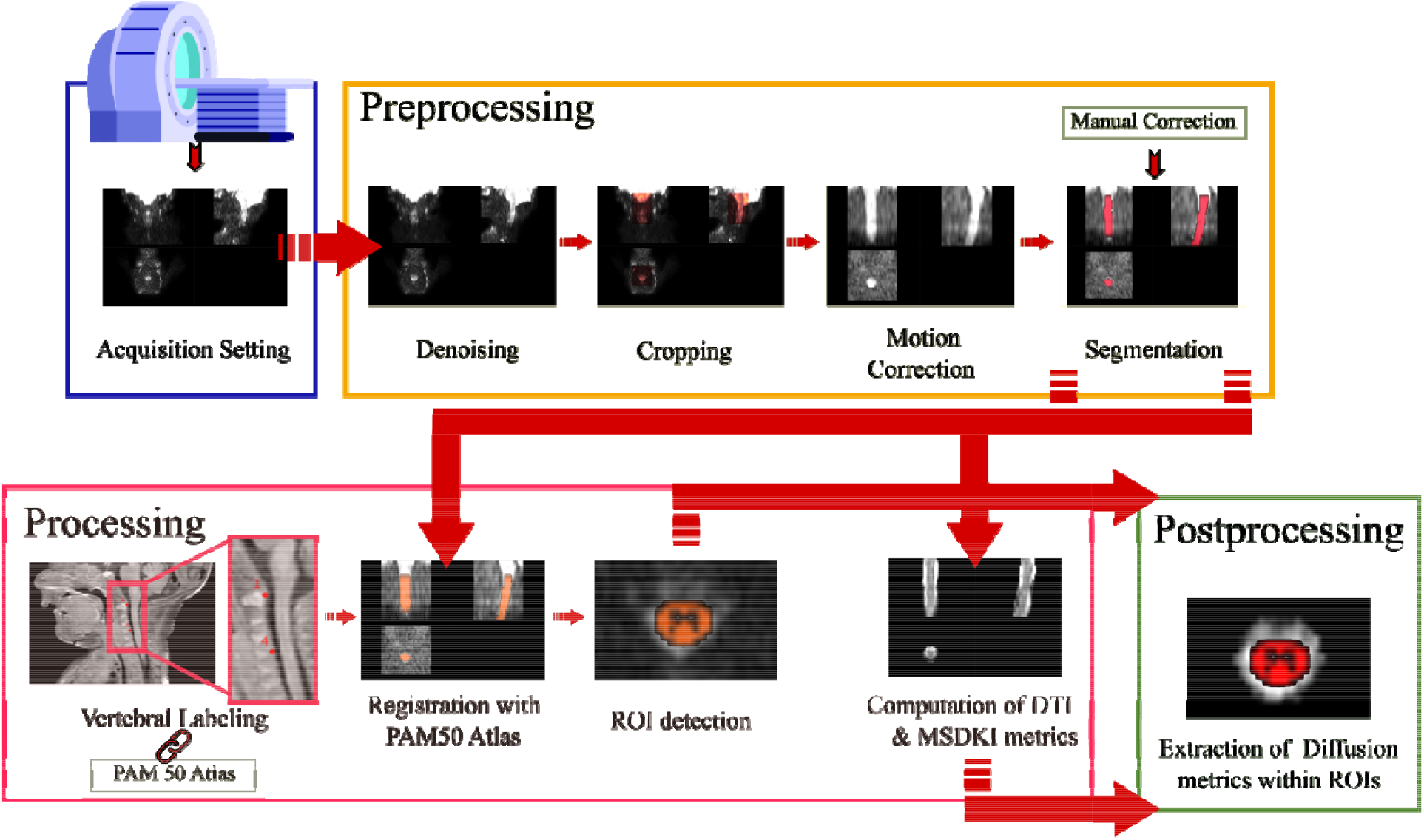
Overall Processing Pipeline: designed pipeline allows complete handling of DKI scan of neonatal Spinal Cord from acquisition setup to preprocessing, processing and postprocessing

### 2.3. Customized acquisition setting

In order to minimize macroscopic movement artifacts, all recommended guidelines for pediatric imaging have been adopted. So as to protect infants from acoustic disturbances caused by MR sequences, we resorted to baby earmuffs and silicone paste for hearing aids. Furthermore, we avoided most of motion by swaddling infants and by placing airbags around the baby’s head. In addition, protective pads have been placed between the magnet and the patient. All these contribute to create a comfortable and warm rest environment, minimizing the chance of free movements.

MRI was performed when possible during spontaneous sleep by exploiting the administration of breast milk or formula about 30 minutes before the start of the exam. In case of spontaneous sleep failure, in order to minimize macroscopic movement artifacts, the instrumental examination was performed under mild sedation by orally administering Midazolam at 0.1-0.2 mg/kg diluted in glucose solution 33%, subject to signature of informed consent from parents and applied by expert trained nurses.

Given the lack of a specific acquisition protocol for DKI of neonatal SC, we designed the diffusion-weighting scheme in collaboration with the neuroradiologists at Giannina Gaslini Hospital. One constraint we had to deal with was the impossibility to perform optimized variants of Spin-Echo Echo Planar Imaging (SE-EPI) sequence (i.e. reduced FOV or spatially selective techniques) (20) on Philips Ingenia scanner. Therefore, minimization of scan duration was our main focus in order to suppress motion and fast CSF pulsation artifacts typical of newborns.

We thus tested different versions of diffusion-weighted gradient scheme, adopting optimal trade-off between Fiber Orientation Distributions profile (FODs, estimated with Mrtrix3 using Multi-Shell Multi-Tissue Constrained Spherical Deconvolution (MS-MT CSD)), image quality and scan time.

We generated each multi-shell diffusion gradient table through Mrtrix3 script *gen_scheme*, taking as inputs the number of phase-encoding directions to be included in the scheme (for most scanners, including ours, typically 1), the b-value of the shell, and the number of directions to include in the shell. This procedure ensures uniform sampling by maximizing uniformity within shells using a bipolar electrostatic repulsion model for optimal angular coverage.

For further reducing acquisition time without significantly affecting image quality, we applied the MultiBand slice acceleration technique (64) (https://www.usa.philips.com/healthcare/resources/landing/compressed-sensecombined).

Moreover, spatial saturation bands have been used to suppress strong signals originating from the subcutaneous lipids to enable volumetric region coverage.

The final version of diffusion acquisition scheme is displayed in Figure 2 as well as reported in Table 2, and includes 6 b=0; 13 b=700 and 13 b=2100 s/mm^2^ for a duration of 4 minutes 30 seconds. This allowed acquisition of high in-plane resolution axial diffusion weighted images, where b=0 scans could be well discriminated from non b=0 volumes and anatomical SC features are sharp.

**Figure 2.**
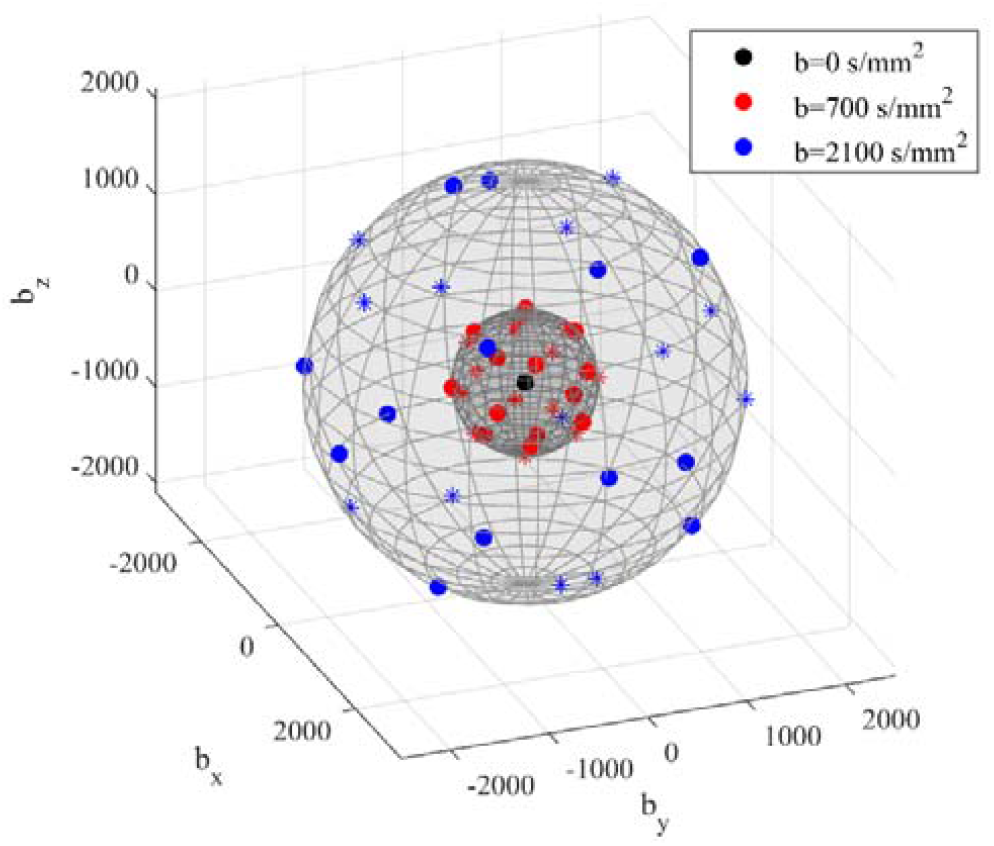
3D view of final diffusion acquisition scheme: directions of diffusion-sensitizing gradients relative to each b-value are displayed in three different colors as reported in the legend. Units are in s/mm^2^. Markers indicates polarity: dots are the polarities in the set, asterisks their opposite Along with dMRI, we also acquired a high-resolution structural image as anatomical reference. The definitive MRI protocol thus consisted in a Turbo Spin Echo (TSE) 3D T1-weighted image followed by a DKI series whose details are listed in Table 1.

**Table 2:**
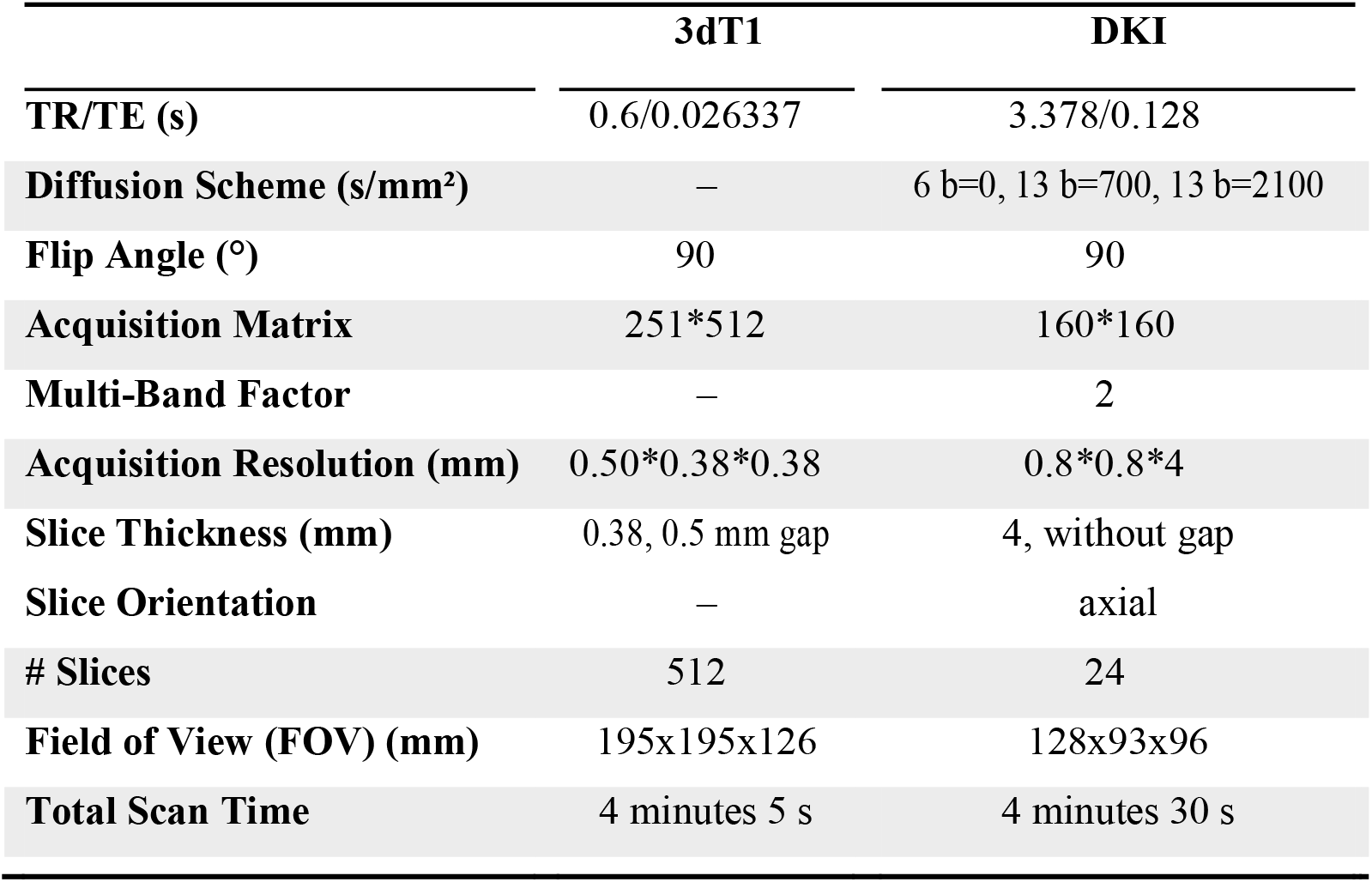
Data Acquisition Details for both structural 3D T1 and DKI image

This is the best possible solution that combines conflicting requirements of complex multi-shell acquisition and minimized scan time.

### 2.4. Preprocessing

#### 2.4.1. Denoising

SC imaging is characterized by low SNR, which can hamper accurate, repeatable, quantitative measurements. Moreover, models such as DKI are susceptible to noise and signal fluctuations often leading to degeneracies in estimation of derived parameters. SNR further lowers in case of neonates, due to relatively high overall free water content, and denoising approaches based on Principal Component Analysis (PCA) are inapplicable due to a reduced number of diffusion gradient directions.

Therefore, we adopted Patch2Self, a recently proposed self-supervised learning denoising method that outperforms existing non supervised methods. (65).

A unique advantage of Patch2Self is the lack of requirement for selecting or calibrating an explicit model either for noise or diffusion signal, so that it can be applied at any step in the pre-processing pipeline. The only assumption it relies on is randomness and uncorrelation of noise across different gradient directions. Its framework consists in holding out one volume and using patches from all other volumes to predict the center of the patches of the held-out volume using a regressor. This denoiser has already showed a significant improvement in repeatability and conspicuity of pathology in diffusion volumes and quantitative DTI metrics for adult SC (66).

Here, we chose to apply Patch2Self as first preprocessing step on raw data since it showed to offer highest SNR. The method is implemented in Dipy v.1.4.0 and applied with Ordinary Least Squares (OLS) regressor, since recommended for SC imaging (Figure 3).

**Figure 3.**
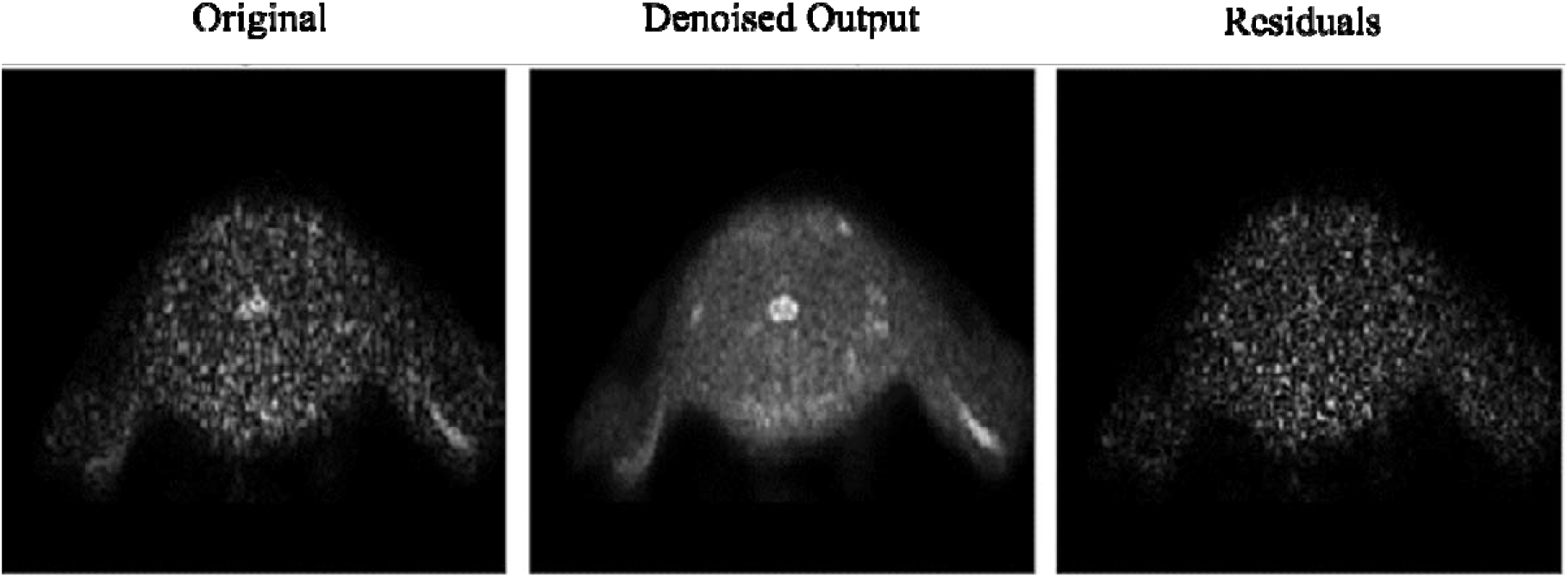
Visual inspection of denoising: The denoising of Patch2Self is compared against the original noisy image along with their corresponding residuals. Notice that Patch2Self does not show any anatomical structure in the corresponding residual plots likely neither introducing structural artifacts

#### 2.4.2. Cropping

SC scans usually include also cerebral areas such as medulla and cerebellum due to their proximity with cervical SC. In order to exclusively focus on the area of interest excluding undesired voxels, as a first preprocessing step, we thus recommend to apply to DKI images SCT function *sct_crop_image* allowing also to fasten subsequent processing. Lower and higher bounds for cropping along the three spatial coordinates can be specified via command line in order to select the same area of interest (i.e. cSC) for all the cohort, considering FOV positioning is consistent across subjects.

Specifically, in the case of our scans, FOV reduction allowed to exclude upper non-spinal areas (i.e. cerebellum) as well as lower spinal levels whose corresponding slices are not usable due to poor image quality (Figure 4a).

**Figure 4.**
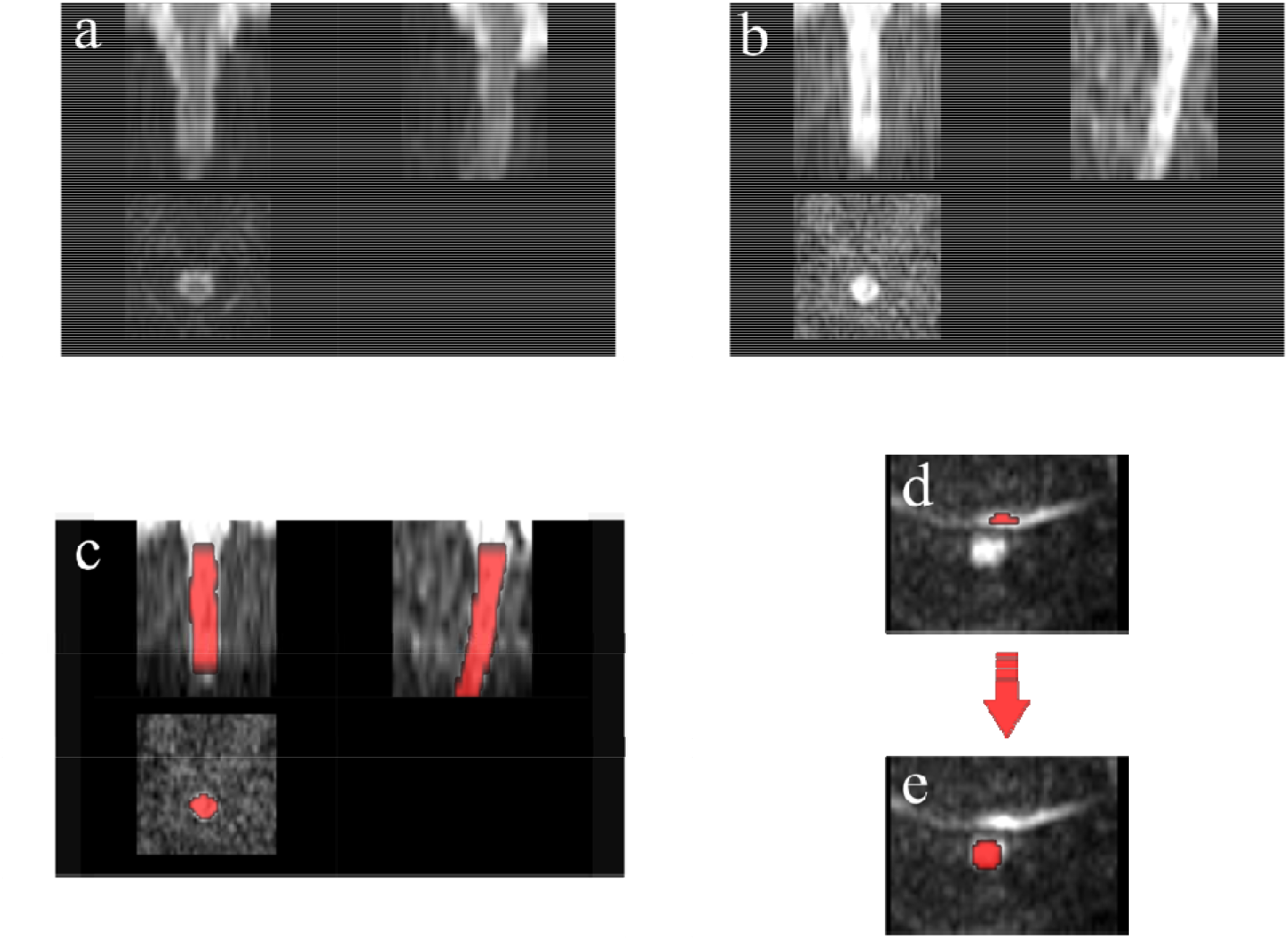
Preprocessing: DKI scan through preprocessing steps for one example subject: (a) FOV reduction; (b) Motion Correction; (c) Segmentation: Deep Learning Segmentation algorithm generally achieves satisfactory results in SC detection; (d) Example of artefactual slice due to a poor fat saturation, causing the fat to alias on the spinal cord area and (e) requiring manual correction of segmentation

#### 2.4.3. Motion Correction

Subjects’ immobilization and anesthesia successfully minimized motion in our acquisitions. However, since dMRI data are analyzed at a voxel level, residual intrascan and/or interslice motion can adversely affect accuracy of the modeled results. We thus resorted to SCT complex motion correction framework *sct_dmri_moco*, based on a combination of tools.

First of all, *SliceReg* algorithm estimates slice-by-slice translations while ensuring regularization constraints along z axis. The latter is achieved using a polynomial function (order specified by the user, flag *-param*). This method was shown to offer better accuracy and robustness than rigid-body transformations and non-regularized slice-by-slice registration, respectively (40).

Moreover, motion correction in SCT includes another feature first proposed in (67) to improve the robustness of registration in high b-value diffusion MRI data such as DKI datasets. It consists in grouping adjacent volumes and estimating the transformation relying on these successive subsets (typically from 3 to 5 volumes) averaged together (flag *-g*).

This robust slice- and group-wise motion correction works successfully also in case of neonatal scans and it is hence applied here with default parameters: grouping of 3 successive dMRI volumes, regularization with 2^nd^ order polynomial function, unitary smoothing kernel (1 mm), and final spline interpolation (flag -x), with the exception of the metric used for registration (Figure 4b). Indeed, Cross Correlation (CC) has been selected as similarity metric given its better performance with respect to Mean Squares or Mutual Information (default option) at the expense of computational time.

Since *sct_dmri_moco* works through iterative average over groups of successive slices in order to increase the SNR of the target image, its output includes a 3D volume corresponding to the mean from DKI slices. These motion-corrected average DKI data will serve as input for subsequent segmentation thanks to its excellent cord contrast.

Thanks to the limited duration of our acquisition and to adopted procedures for minimizing movement throughout the exam, amount of motion is very limited in our images. As a result, outcome of motion correction step does not significantly differ from raw DKI image by visual assessment. However, this represents a crucial step in case of longer scans more prone to source of motion artefacts.

#### 2.4.4. Segmentation

Proper segmentation of SC is decisive for subsequent steps of template registration and computation of metrics along the cord.

Detection of SC has turned out to be a critical step, since standard SCT algorithm *propseg*, based on multi-resolution propagation of tubular deformable models (68), is trained for adult spine.

Given the reduced size of neonatal SC and the low contrast between the spine and CSF, default segmentation method fails in several slices even after modulating the algorithm parameters - e.g. manual initialization of spinal cord centerline through interactive viewer (flag *-init-mask*), selection of SC radius size (flag *-radius*) or cord rescale (flag *-rescale*).

We thus resort to a more recent and advanced method of SC extraction, based on deep learning *sct_deepseg_sc* (69). This fully automatic segmentation framework was conceived for detecting SC and intramedullary MS lesions from a variety of MRI contrasts and resolutions.

It is composed of a cascade of two convolutional neural networks (CNN), specifically designed to deal with spinal cord morphometry: the first detects the cord centerline and reduces the space around the spinal cord (for better class balance), and the second segments the cord.

Segmentation results outperformed *sct_propseg*, showing higher robustness to variability in both image parameters and clinical conditions.

Thanks to its versatility, the application of this method results suitable also for neonatal imaging, allowing robust and accurate segmentation of our scans without ever the need of additional parameters but just specifying the kind of image contrast as *<u>dwi</u>* (flag *-c*) (Figure 4c).

In case of failure of SC detection, we necessarily opt for manual correction of problematic slices on FSL editor (*FSLeyes*) (Figure 4d,e).

This is the case of five subjects within our cohort: to validate the quality of segmentation, we checked the QC feature on our MRI images across subjects and noticed some local segmentation leakage - (related to the onset of artifacts at acquisition phase and not to a flaw with the algorithm) - in a few slices and hence corrected it manually.

### 2.5. Processing

#### 2.5.1. Vertebral Labeling

After segmentation, labeling of vertebral levels or discs is the second mandatory step in order to match the template to the subject’s MRI (template registration).

Two vertebral levels are necessary for registering data to the template. Each of these two landmarks consists of a voxel placed in the middle of the SC, at the level of the corresponding mid-vertebral body, and assigned a relative number starting from 1 for C1 vertebra. However, SCT recently introduced the possibility to alternatively use inter-vertebral disc labels with the analogous procedure of reference numbered voxels.

We perform this step on 3D T1w images in order to achieve better accuracy given their higher overall quality and contrast compared to DKI ones, where vertebral discs are not clearly identifiable.

Labeling from 3D T1w anatomical image is possible as it turned out to match relatively well along the superior-inferior (z) axis, the target direction of disc labeling, with the DKI scan (not along the Anterior-Posterior (AP) or Right-Left (RL) direction, Figure 5).

**Figure 5.**
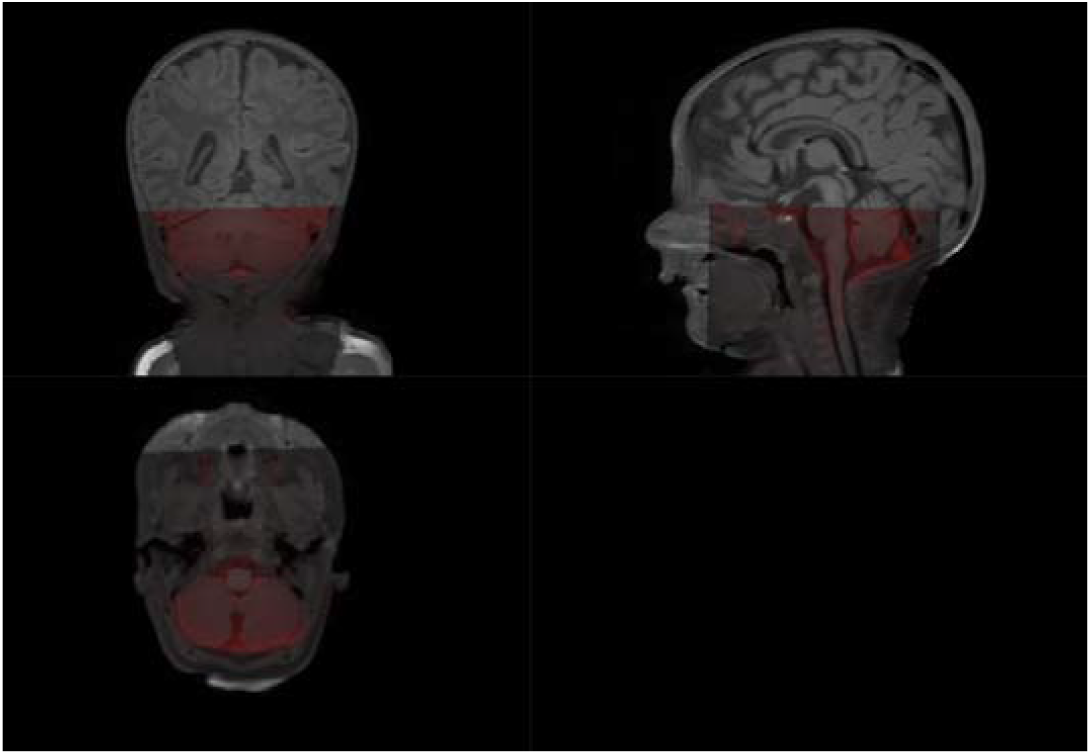
DKI scan overlaid on structural 3dT1w image: while both images are clearly not registered along the antero-posterior direction due to the very strong susceptibility artefact, the z-location is similar: see how the bottom tip of the cerebellum is consistent for the two scans

Vertebral labeling is typically done using an automatic method *sct_label_vertebrae*, that finds C2-C3 disc, and then locates neighbouring discs using similarity measure with the PAM50 template at each specific level (70).

Default SCT procedure *sct_label_vertebrae* fails in automatically detecting C2-C3 vertebral disc once again because of the small size of spines at issue and low image contrast compared to adults.

Therefore, we manually create labels with the command *sct_label_utils* through interactive viewer option provided by SCT (flag *-create-viewer*) with little to no waste of time.

Specifically, vertebral labeling was created at the posterior tip of the top of C1 vertebra and at C3-C4 disc, centered in the cord. Manual intervention only took a few seconds per subject (Figure 3S).

#### 2.5.2. Registration to PAM50 atlas

Registration between subject’s diffusion and atlas space is a very demanding task in case of neonatal imaging given the lack of a specific pediatric atlas compatible with SCT (one is currently under creation, https://github.com/neuropoly/spinalcordtoolbox/issues/2530). We thus use PAM50 atlas (71), an adult template for MRI of the full SC and brainstem in the same coordinate system as the ICBM152 (MNI) brain template, allowing to conduct simultaneous brain/spine studies. It consists of a T1w, T2w, T2*w, white and gray matter probabilistic atlas and white matter atlas of tracts as well as probabilistic labeling of spinal levels. The template has been constructed from straightened SC for facilitating registration and visualization of results.

*sct_register_to_template* is the main command for registering one subject to the template and vice versa, since it outputs the forward and backward warping fields. We choose subject’s native diffusion space as target of registration transforms since the straightening required by opposite strategy would cause through-plane interpolation errors which would bias following extraction of diffusion measures (72).

Moreover, we suggest employing T1w atlas image for its better contrast similarity with DKI scan compared to T2w.

Application of default command does not produce satisfactory results, stressing the need to tweak all input parameters to deal with our particular contrast and resolution. Given the presence of artifacts and some inherent features (e.g., low CSF/cord contrast) that could compromise the registration, we use SC segmentation as input for the algorithm in order to ensure maximum robustness.

Registration is then built through multiple steps by increasing the complexity of the transformation performed in each step (starting with large deformation with low degree of freedom and finishing with local adjustment). Specifically, the first step consists in vertebral alignment, that is vertebral level matching between the subject and the template on the basis of posterior edge of the intervertebral discs provided by previous manual vertebral labeling. Second step is slice-wise center of mass alignment between the two images, using *centermass* algorithm instead of default *centermassrot* (which also includes rotation alignment) because the cord is quasi-circular and cord angle estimation is not reliable here. The third step is R-L scaling along x axis followed by A-P alignment to match segmentation borders along y axis, with the ultimate aim of accommodating the very small SC size. Finally, iterative slice-wise non-linear registration is performed through non-linear symmetric normalization regularized with b-splines (73) using information from comparison of Cross Correlation metric (CC) between the two images, which allows SC shape refinement. Once the algorithm completed, one can assess the quality of registration through visual evaluation and inspection of QC module, and thus warp the template and all its objects to each subject’s DKI image (Figure 6).

**Figure 6.**
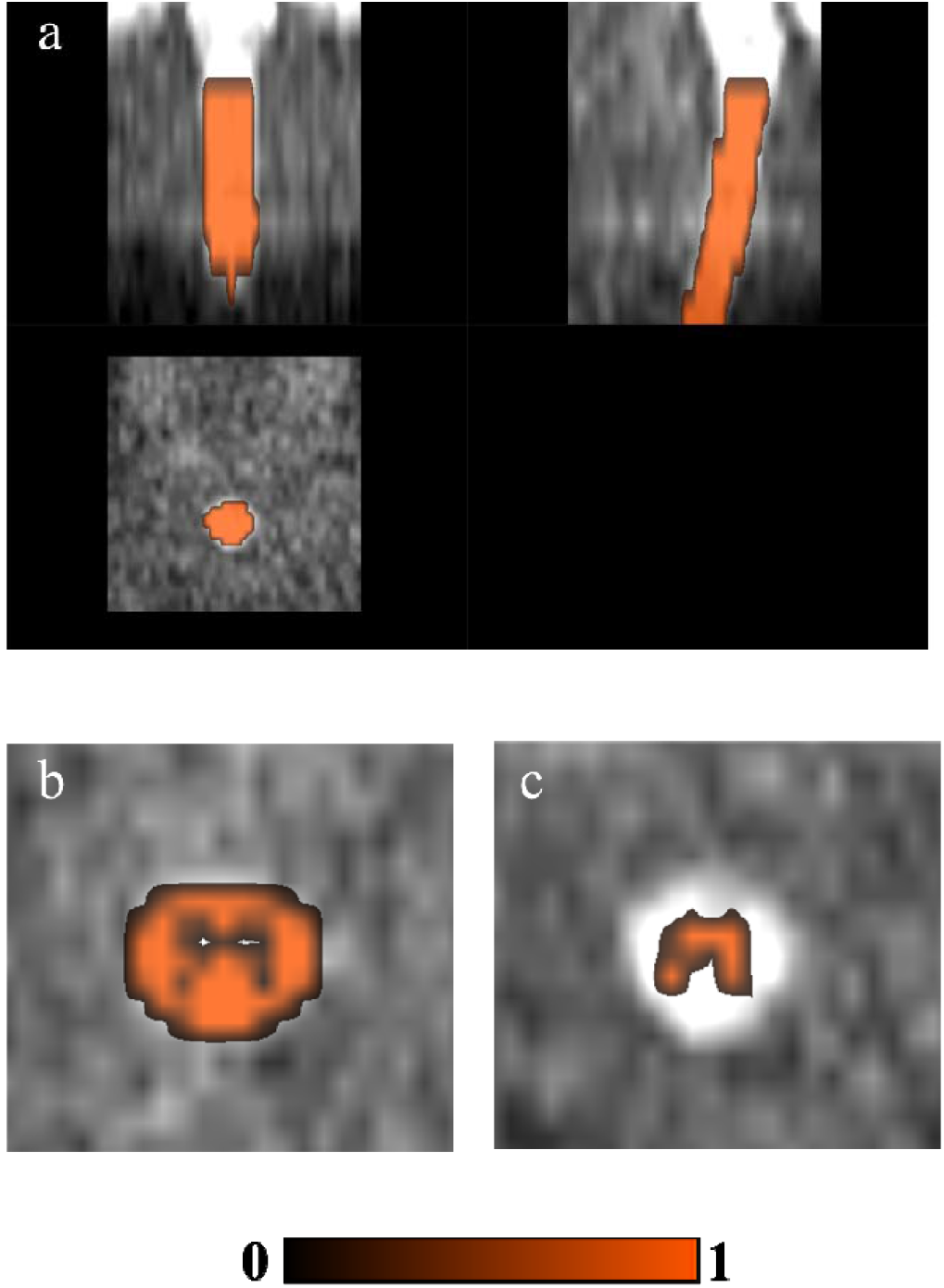
Registration with PAM50 atlas and ROI detection through atlas-based approach: (a) PAM50 atlas’ cord segmentation binary mask, (b) WM and (c) GM probabilistic masks warped to subject’s DKI motion-corrected mean image

Current selection of parameters and steps successfully worked for our scans since atlas registration algorithm robustly achieved convergence as verified through inspection of QC feature.

#### 2.5.3. Computation of diffusion metrics

The end point of previous preprocessing and processing steps is computation of diffusion parametric maps, from which to extract quantitative summary measures requested by the particular study in question. We estimate diffusion parametric maps through DIPY software (v. 1.4.0), (41) To avoid unnecessary calculations on the background of the image, we use a mask created by dilating spinal cord segmentation (through *sct_maths* command) because values outside the binary cord mask are important for proper account of PVE, having to be minimized in every possible way (74). Indeed, this phenomenon, because of the coarse resolution of MRI with respect to SC anatomy, may make the apparent value within a boundary voxel be a mixture between the WM and CSF compartment, thus yielding a subsequent inaccurate quantification of diffusion measures.

Since DKI model involves the estimation of a large number of parameters (75) and is more sensitive to artefacts (76), we choose to further suppress the effects of noise and artefacts before diffusion kurtosis fitting using a 3D Gaussian smoothing (with a Gaussian kernel with fwhm=1.25) as suggested by pioneer DKI studies (2). This also helps in addressing the issue of implausible negative values inherent to DKI fitting (77).

The following parametric maps can thus be generated: Mean Diffusivity (MD), Axial Diffusivity (AD), Radial Diffusivity (RD), Fractional Anisotropy (FA) and Mean Kurtosis (MK), Axial Kurtosis (AK), Radial Kurtosis (RK), Kurtosis Fractional Anisotropy (KFA), and Mean Signal Kurtosis (MSK).

Given the low-angular resolution data available, to ensure robustness and reproducibility of parameters’ estimates, we opted for just computing DTI measures, whose reference tensor can be correctly estimated from at least six independent directions, and MSK, as displayed in Figure 1S. The latter is a robust scalar kurtosis index that can be estimated from powder averaged diffusion-weighted data and thus independent from acquisition scheme (78,79). Indeed, fitting MSDKI is well posed without relying on the full DK tensor, which would require a minimum of 15 non-collinear directions per b-value. Moreover, this measure is generally more robust to low SNR situations as in case of neonatal imaging.

This novel DKI estimator can be seen as a proxy for the MK, showing to present nearly identical contrast to the original DK metric while improving robustness and reproducibility of the kurtosis metrics and results in parameter maps with enhanced quality and contrast. Specifically, this measure turns out to be less sensitive to thermal noise and imaging artifacts, and thus drastically reduces black voxels intrinsic to DKI and challenging the visual and statistical analysis of potentially clinically relevant biomarkers of tissue integrity. Moreover, as previously pointed (80), standard kurtosis measures do not only depend on microstructural properties but also on mesoscopic properties such as fiber dispersion or the intersection angle of crossing fibers. In contrary, the kurtosis from powder-average signals has the advantage of being decoupled from confounding effects of tissue dispersion and crossing (80,81).

### 2.6 Postprocessing

Thanks to this atlas-based analysis approach, it is possible to perform cord-specific quantification of diffusion metrics through *sct_extract_metric* command, also restricted to specific Regions of Interest (ROIs; labels used by default are taken from the PAM50 template, e.g. WM tracts, flag *-l*), vertebral levels (flag *-vert*) or slice (flag *-z*), according to the specific clinical needs concerned.

Along with WM and GM probabilistic masks as a whole (Figure 6b,c), normally investigated in medical practice, one can carry out ROI detection also in specific tracts according to the clinical question (fifteen WM tracts and three GM regions available in total for each side).

In our example, neither DKI nor structural images ensured sufficient WM-GM-CSF contrast to perform any manual detection of ROIs in contrast to high-contrast PSIR image of (82), whose acquisition time would be too long for neonates. Therefore, we exploited good registration outcome for automatic delineation of ROIs through atlas-based approach.

We opted for using lateral CSTs as ROIs for consistency with (82) - though grouping together left and right sides in order to gain robustness by increasing volume fraction as suggested in (40) - as well as WM and GM.

We then computed averages of diffusion measures across C1-C4 vertebral levels in order to avoid a subject-based bias: outside of these levels the registration is inaccurate and/or MRI signal may be corrupted. We thus checked through QC module the correctly segmented slices corresponded to the same vertebral levels across subjects, starting from the first slice containing only SC (excluding cerebellum, Figure 2Sc).

Moreover, estimation of DTI and MSDKI weighted average metrics was limited to those slices where spinal cord segmentation is accurate: outside segmentation mask, metrics would indeed be irrelevant. This was obtained by multiplying segmentation mask by specific WM, GM and CSTs atlas labels. We quantified diffusion metrics using Weighted Average (WA) estimation to minimize PVE avoiding bias into resulting metrics by the surrounding tissues (e.g. CSF). This is one of the recommended methods especially in case of noisy images and small tracts as in our case. We assessed associated voxel fraction to quantify the reliability of our diffusion measures: as demonstrated in (40), having at least 240 voxels results in an error smaller than 1%, while having 30 voxels results in an error inferior than 2%. In this example, the metrics were computed based on average 178.3, 50.5 and 31.5 voxels in WM, GM and CSTs respectively, thus assuring sufficient accuracy of estimates.

### 2.7 Case Study

Periventricular WM Injury (PMWI) is the most frequent type of brain lesion in preterm infants, and the spatial extent and location of WM injury correlate with distinct clinical outcomes, including cerebral palsy and motor impairment (83).

Given the strong association of WM injury with the motor function development of preterm neonates, we assessed the hypothesis that the presence of periventricular punctate WM lesions identified on MRI at TEA could be associated with regionally specific alterations in cSC microstructure.

A similar approach was already used by (82) to characterize cSC microstructural abnormalities in a cohort of adult patients with previous unilateral ischemic stroke in the vascular territory of the middle cerebral artery (MCA). DTI and DKI diffusion measures in cSC resulted to be a valuable imaging marker for predicting clinical outcome. In particular, significant reduction of FA and MK was observed in the affected lateral WM bundle of the cSC, correlating with the severity of motor dysfunction.

Accordingly, the ultimate goal of our study was to verify whether the presence of periventricular WM lesions affects the cSC tracts development. Specifically, we aimed to compare DTI and MSDKI measures of cSC in two groups of preterm neonates: (i) with punctate Periventricular White Matter lesions (PWMI), and (ii) with normal brain MRI (controls).

## 3. Results

### 3.1. Population size and classification

In order to investigate clinical differences among acquired subjects, we grouped infants as follows: i) 9 subjects with punctate PWMI and ii) 9 subjects with normal brain MRI, used as control group. At QC phase, in accordance with the expert neuroradiologist, we opted for excluding one control subject due to excessively poor image quality (i.e. signal leakage at C1-C3 level, Figure 2Sa,b). Therefore, the final number of subjects under analysis amounted to 9 and 8 infants for the two subsets, respectively.

### 3.2. The role of denoising

As mentioned above, neonatal imaging is inherently affected by low SNR and sensitive to imaging artifacts. Proper denoising of scans is therefore a crucial step in the processing pipeline. Above all, we thus focused on quantitatively assessing the contribution of Patch2Self denoiser on subsequent analysis.

Firstly, we computed average SNR on b=0 images, namely thermal noise due to the diffusion-related signal attenuation, across all slices belonging to C1-C4 district of our interest.

As expected, we found a remarkable increase in mean SNR after applying Patch2Self (5.88 ± 1.41 vs 14.64 ± 4.53) as displayed in Figure 7.

**Figure 7.**
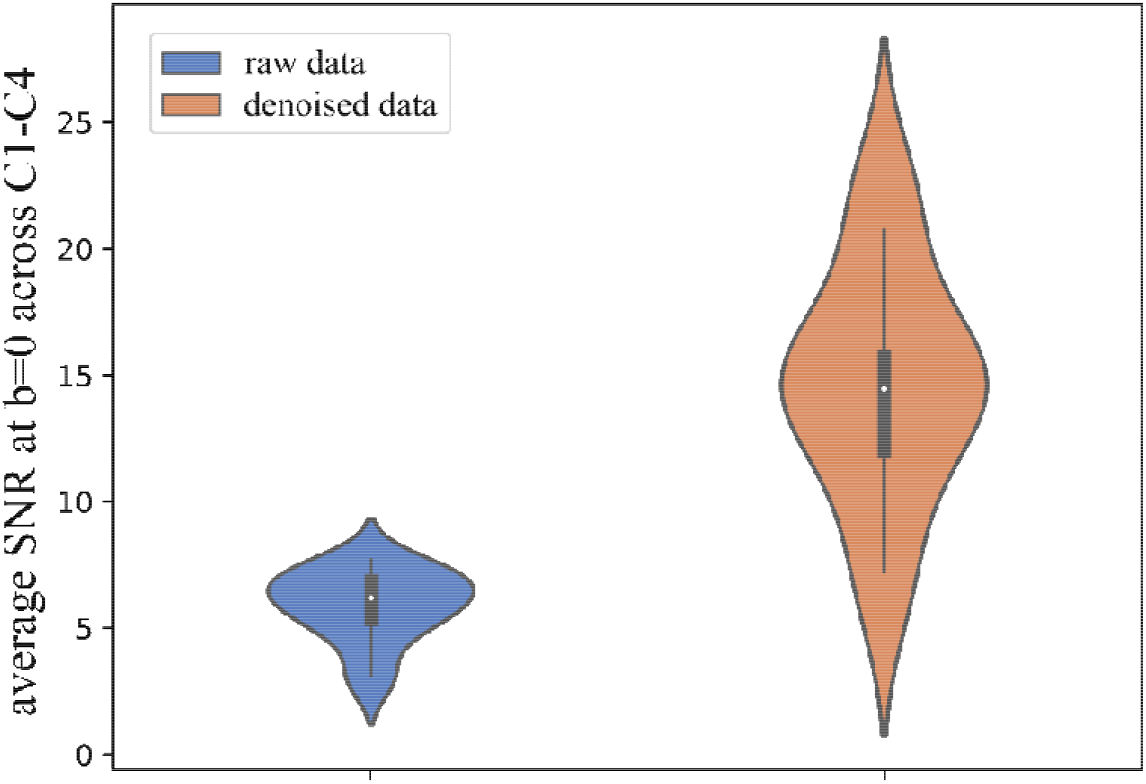
Effects of Patch2Self denoising on thermal noise. Average Signal-to-Noise Ratio (SNR) computed on b=0 images increases, across C1-C4 vertebral levels under analysis, when including denoising with Patch2Self algorithm in the processing pipeline

We then inspected impact of denoising on microstructure model fitting, a critical step often leading to degenerate parameter estimates due to the low SNR of dMRI acquisitions.

Specifically, we applied DTI and MSDKI model on raw and denoised data, resorting both to traditional Marchenko-Pastur PCA (MP-PCA) and to Patch2Self method. We opted for comparing our denoising procedure with MP-PCA since it represents the current state-of-the-art unsupervised method for denoising DWI (84). In order to compare the goodness of each fit, we performed a k-fold cross-validation (85) across an axial slice (1312 voxels) of masked data for an example subject pulled out from our dataset. In Figure 8, we depict the improvement of the metric by simply subtracting the goodness-of-fit scores of fitting noisy data from Marchenko-Pastur and Patch2Self denoised data for both DTI and MSDKI models. Patch2Self performance is not significantly better than MP-PCA in the goodness-of-fit for DTI model (two-sided t-test with Bonferroni correction, p=1). On the contrary, this state-of-the-art denoiser shows a significant improvement for MSDKI (p=1.825e-04), proving once more to be particularly suitable for DKI model.

**Figure 8.**
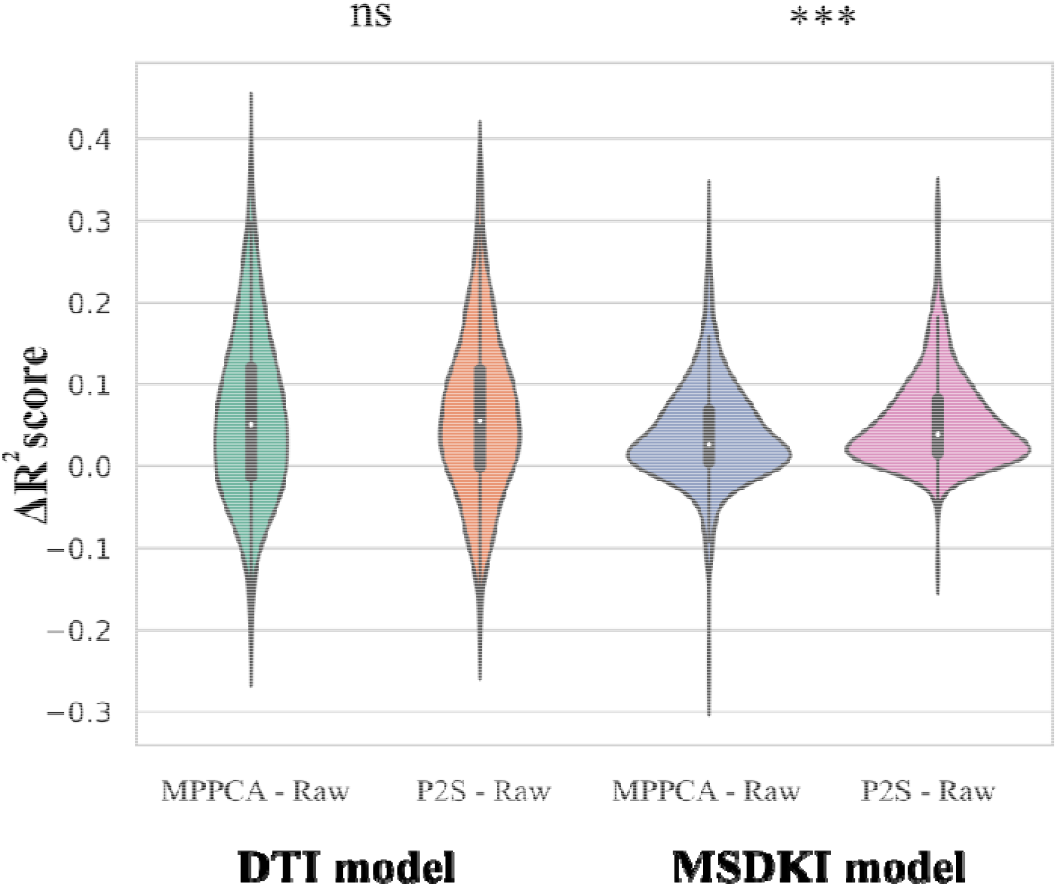
Violin plots quantifying the increase in metric after fitting downstream DTI and MSDKI models for one axial slice in one example subject. The improvements in each case are plotted by subtracting the scores of model fitting on noisy data (Raw) from of fitting each denoised output. Note that the consistency of microstructure model fitting on Patch2Self (P2S) denoised data is higher than that obtained from Marchenko-Pastur (MPPCA), especially as regards MSDKI model. ns stands for non significant, *** stands for 1.00e-04 < p <= 1.00e-03 in two-sided t-test with Bonferroni correction

### 3.3. Trend of diffusion measures

For consistency with (82), we considered above all trend of cSC FA and MSK measures in the two groups of neonates as displayed in Figure 9. For the sake of completeness, in Figure 4S we also include remaining DTI metrics.

**Figure 9.**
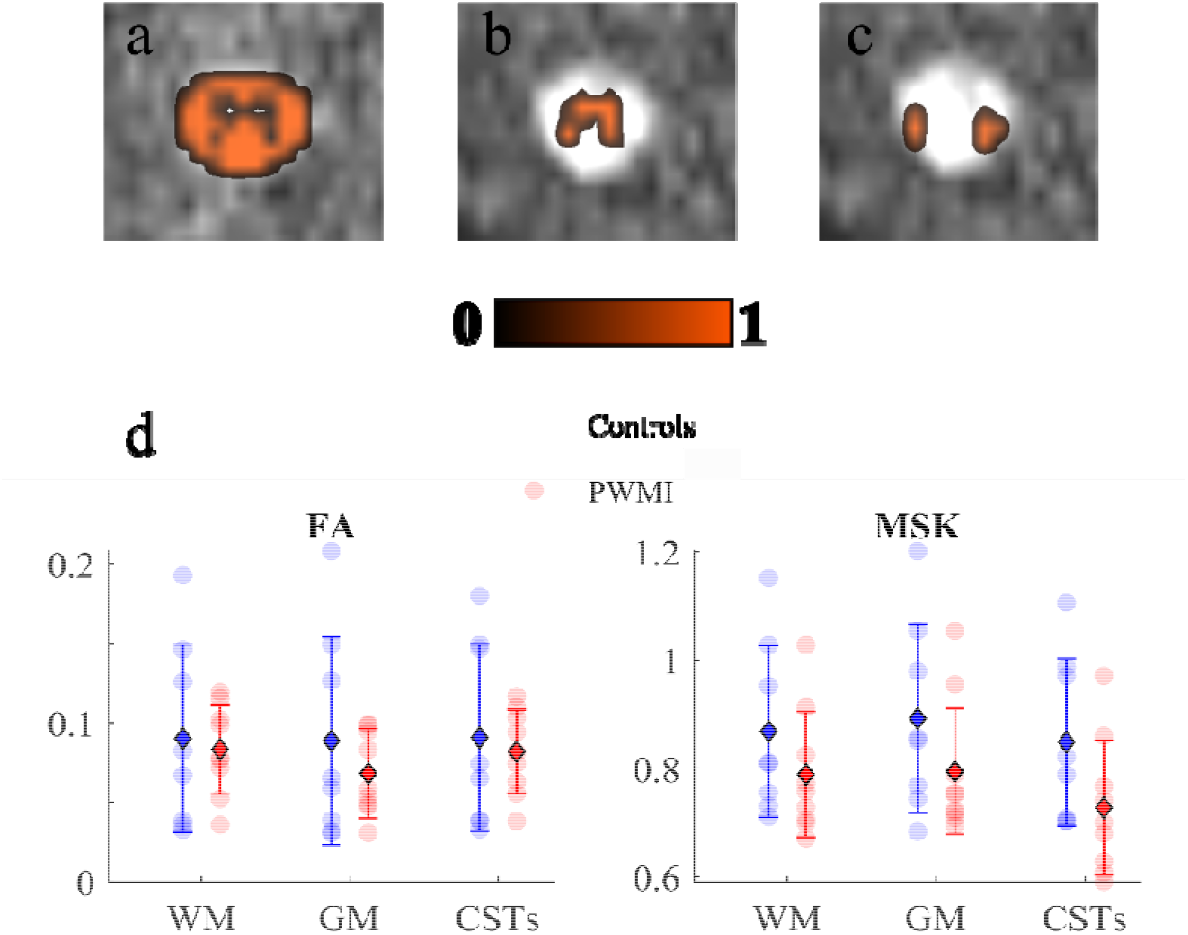
Extraction of diffusion measures within specific ROIs: (a) White Matter (WM), (b) Gray Matter (GM) and (c) Cortico-Spinal Tracts (CSTs) ROIs overlaid on DKI motion corrected image; (d) Scatter plots of FA and MSK in group subjects across aforementioned ROIs: colored spots indicate single subject’s value for each metric; as reported in the legend, controls’ measures are in blue, whereas Periventricular White Matter Injury (PWMI) group’s in red. Units for MSK are in mm^2^/s, while FA is dimensionless. Error bars displaying mean (diamond) and standard deviation (bars) are overlaid on scatter plots.

To an initial evaluation based on the limited sample size available, considering the variation of average metrics between controls and patients in WM, we can notice an increase in MD, AD, RD, parallel to an overall decrease in FA and MSK in preterm neonates with PWMI.

Furthermore, range of values of DTI measures are consistent with normative values on healthy pediatric SC (30). MD, AD and RD values are higher while FA values are lower compared to equivalent measures on older cohorts of patients (i.e. children/ adolescents, (22,23,25,26)). This may be partially due to sensitivity of FA to denoising, which can imply a reduction in this metric. Moreover, this trend is in line with the simultaneous age-related decrease in MD, AD and RD and increase in FA metrics reflecting progressive maturation, myelination and fiber packing and thickening within the SC, similar to that observed in the brain (86,87).

Conversely, definition of a normative variation of DKI measures across age from newborns to adults will be feasible after further investigations from early stages of development.

## 4. Discussion

### 4.1. Research question

In the present study we tested the pipeline of SC DKI analysis in a group of neonates with PWMI, a form of mild WM injury frequently diagnosed in preterm infants. PWMI are seen at brain MRI as small, focal, multiple alterations of signal intensity (high on T1 and/or low on T2) in periventricular WM. Long-term outcome of neonates with PWMI seems to be related to the number of lesions, their pattern, and their localization. Of note, several studies have showed that greater lesion load of PWM and the involvement of frontal WM are associated with higher risk of adverse neurodevelopmental outcome, affecting both motor and cognitive functions (88). Moreover, periventricular WM lesions in preterm neonates are associated with region-specific changes in MD, FA, RD, and AD in several cerebral WM tracts that might explain the abnormal development of long-term neurological functions (89). Specifically, the involvement of pyramidal tract fibers in the periventricular WM has been demonstrated to be a relevant factor for motor dysfunction in children with PWMI (90). In our study, we found that microstructural changes can be detected by using an advanced DKI analysis also in the GM and WM of cSC of preterm neonates with PWMI studied at term-equivalent age, thus demonstrating that DKI parameters could be used as markers to unravel underlying microstructural lesions not visible on conventional structural MRI. Moreover, our preliminary findings confirm the hypothesis that in preterm neonates with PWMI WM microstructure alterations extend beyond the immediate area of periventricular injury, widening distally also in the cSC (91,92). Further analyses on a wider cohort of neonates are necessary to confirm these preliminary results, and specifically to prove if microstructural changes in cSC in preterm neonates with PWMI correlate with long-term neurological outcome.

### 4.2. Study significance

Here, we present the first application of DKI to neonatal SC through a pipeline able to perform complete processing on a subject within a clinically acceptable time (10 minutes average with the current setup). As regards acquisition setting, we were able to perform a time-consuming technique like DKI using a short diffusion sequence which minimizes patient’s physiological motion and which likely reflects a standard clinical scenario devoid of latest technologies in terms of acquisition sequence optimization. With regard to image processing, we opted for creating this pipeline using SCT since it represents the only existing comprehensive, free and open-source software dedicated to the processing and analysis of SC multi-parametric MRI data. Adaptation of each image processing tools already in use for adult subjects through appropriate tuning of parameters turned out to be feasible. Indeed, it allowed to successfully overcome all the issues mentioned in the ‘Introduction’ section inherent to imaging of SC and exacerbated in case of neonatal setting, even for the most challenging steps like segmentation or registration to atlas. We were thus able to quantify diffusion measures within specific ROIs using an atlas-based approach which presents undisputed advantages compared to usual manual drawing of ROIs. It is automatic and thus highly reproducible, it is not biased by the user experience and knowledge of the anatomy, it is much faster than long and tedious manual delineation of ROIs and it allows to account for PVE. Preliminary application of our method to a limited number of subjects has yielded physiologically plausible findings despite suboptimal acquisition setting. In particular, differences in DTI and MSDKI measures observed between PWMI neonates and controls confirm the hypothesis that PWM injuries in the premature brain can be associated with microstructural alterations of both GM and WM of neonatal cSC at term-equivalent age. These findings also highlight the importance of complementary analysis of DTI and DKI metrics for a more accurate characterization of biological tissues.

### 4.3 Added value of DKI

Results about feasibility of DTI and MSDKI analysis in neonatal SC with subjects collected so far are preliminary but promising and demonstrate the clinical utility of combining DTI and DKI in the characterization of spinal cord pathologies.

FA reduction parallel to MD increase in patients is an expected finding consistent with existing literature and attributable to degeneration of diffusion barrier and loss of diffusion directionality.

Regarding DKI, it is interesting to note how MSK metric, although yet underused in clinical studies, proves to be more sensitive than FA to microstructural changes in PWMI subjects by further reducing its value in all considered ROIs.

This DKI estimator thus confirms DKI increased sensitivity in capturing alterations related to pathology, also far from the lesion site.

Another crucial food for thought concerns observing how the presence of a WM lesion in the brain causes subsequent alterations not only in cSC WM but also in GM, as evidence of the strong association between brain and spine. In this respect, resorting to DKI measures becomes of utmost importance given kurtosis sensitivity to structural changes in isotropic tissues such as GM.

Indeed, range of variation of MSDKI metric from controls to PWMI is overall higher in GM than that of corresponding DTI measures.

This is an expected finding: DTI is extremely suitable for capturing highly directional diffusion typical of WM bundles but shows low sensitivity to isotropic diffusion, whereas DKI allows specific assessment of non-gaussian diffusion typical of GM areas (93).

Such findings once again stress the importance of combining DTI and DKI metrics as complementary sensitive biomarkers in order to fully exploit the potential of dMRI compared to conventional MRI.

However, a more comprehensive corroboration and explanation of our results is expected after collecting an adequate number of subjects to carry out a robust statistical survey. An in-depth interpretation of the single metrics is out of the scope of this paper. Here, we just dwell on exploring the comparison with the work on adults which served as a starting point.

### 4.4 Comparison with adult study

Since MSK has proved to be an exact approximation of MK, here we assimilate it with standard DKI metric used in (82).

Both FA and MSK trend agree with just mentioned study. Specifically, they both exhibit a reduction in WM, GM and CSTs in case of pathology (Figure 9).

Since FA is known to be an index of structural integrity (94) and MSK a marker of tissue microstructure complexity (2), obtained findings suggest that, in case of overlying WM brain lesion, a loss of integrity and complexity is registered also in SC WM tracts below with a more isotropic diffusion pattern due to disruption of WM tracts. Also MD, AD and RD follow the same trend, with an increase in case of lesioned subjects, as in (82). This biologically plausible hypothesis already verified for adults would thus also subsist in infants.

### 4.5. Study limitations

Present pipeline has been designed with current instruments at disposal and with given data whose resolution is high in axial plane but whose contrast is very low due to short acquisition time. As a result, any improvement in acquisition setup our pipeline will bring to even stronger and more comprehensive results.

Major flaws of this procedure consist in basing on an adult atlas, where the exact location of tracts may not perfectly correspond to neonatal images despite the good adjustment of registration parameters. Current pipeline will definitely benefit from introduction of a pediatric atlas into SCT. Moreover, image quality could be further improved: scans we acquired are extracted from routine clinical protocol and consequently prone to noise and artifacts due to short acquisition time dictated by clinical needs and to the lack of specific spatially-selective MR sequences.

Starting from more advanced hardware tools may significantly increase image quality and thus accuracy of estimated metrics. Resorting to optimized acquisition sequences would also allow to increase resolution of HARDI acquisition scheme and thus to exploit all standard DKI measures, which can in turn increase the amount of diagnostic information.

We acknowledge the protocol in use to be on the edge for HARDI schemes required by DKI. However, this represents an attempt to customize advanced dMRI acquisition setting within a clinical routine protocol, already long in itself since made up of multiple MRI sequences in order to increase diagnostic possibilities. Nevertheless, we appropriately addressed this issue at DKI tensor and measures computation phase to ensure reliability and accuracy in their estimates.

### 4.6. Future developments

Validation of current pipeline can be made by testing it to a larger cohort of subjects, possibly investigating also lower SC tracts, including thoracic and lumbar districts, and extending studies to different clinical cases, preferably focusing on a determined pathology. For example, it would be interesting to explore long-term correlations between DKI measures and specific clinical scores as done in (82), where diffusion measures have been related to motor performance indexes.

A further step may be adapting this analysis pipeline to other promising higher-order diffusion models requiring multi-shell acquisition such as NODDI (95).

## 5. Conclusion

In this work, we have showed how accurate adjustment and parameters’ tuning of processing algorithms customized for adult SC opens up new horizons in exploiting increased ability of advanced dMRI models, also in neonatal domain, where they had never been utilized before.

Indeed, even starting from low quality data acquired for diagnostic purposes and thus suboptimal, we were able to extract from DKI information of some diagnostic use.

The case study proposed in this paper is just an example of the potential relapses of this semi-automated pipeline, which paves the wave for applying advanced dMRI models to neonatal setting in a wide range of potential clinical applications. In particular, the possibility of successfully exploiting increased sensitivity and sensibility inherent to DKI methodology also into neonatal setting would indeed be extremely useful for throwing light on complex diseases related to this critical phase of development and to deepen the knowledge about the relationship between brain and SC at birth.

## Data Availability

Data used in this study is not able to be made openly available due to privacy restrictions of clinical data imposed by the Gaslini Hospital's administration. Regarding code availability, a specific pipeline compatible with SCT is integrated in an open-access GitHub repository within "SCT-pipeline", a site gathering various pipelines compatible with SCT for processing MRI data (https://github.com/sct-pipeline/pediatric-genova).

## Author contributions

**Rosella Trò:** Conceptualization, Methodology, Software, Writing - Original Draft, Visualization. **Monica Roascio**: Conceptualization, Supervision. **Domenico Tortora**: Conceptualization, Validation, Investigation, Resources, Supervision. **Mariasavina Severino**: Validation, Investigation, Resources, Supervision. **Andrea Rossi:** Validation, Investigation, Resources, Supervision. **Julien Cohen-Adad:** Writing-Reviewing and Editing, Supervision. **Marco Massimo Fato:** Conceptualization, Writing-Reviewing and Editing, Supervision. **Gabriele Arnulfo:** Conceptualization, Writing-Reviewing and Editing, Supervision. **These last two authors contributed equally**.

## Funding sources

This research did not receive any specific grant from funding agencies in the public, commercial, or not-for-profit sectors.

Acknowledgements

The authors would like to thank Prof. Luca Antonio Ramenghi (Neonatal Intensive Care Unit, IRCCS Istituto Giannina Gaslini, Genoa, Italy e Department of Neurosciences, Rehabilitation, Ophthalmology, Genetics, Maternal and Child Health (DINOGMI), University of Genoa, Italy) and the LIFT (Laboratorio di Imaging Funzionale 3 Tesla).

## Supplementary material

**Figure 1S.**
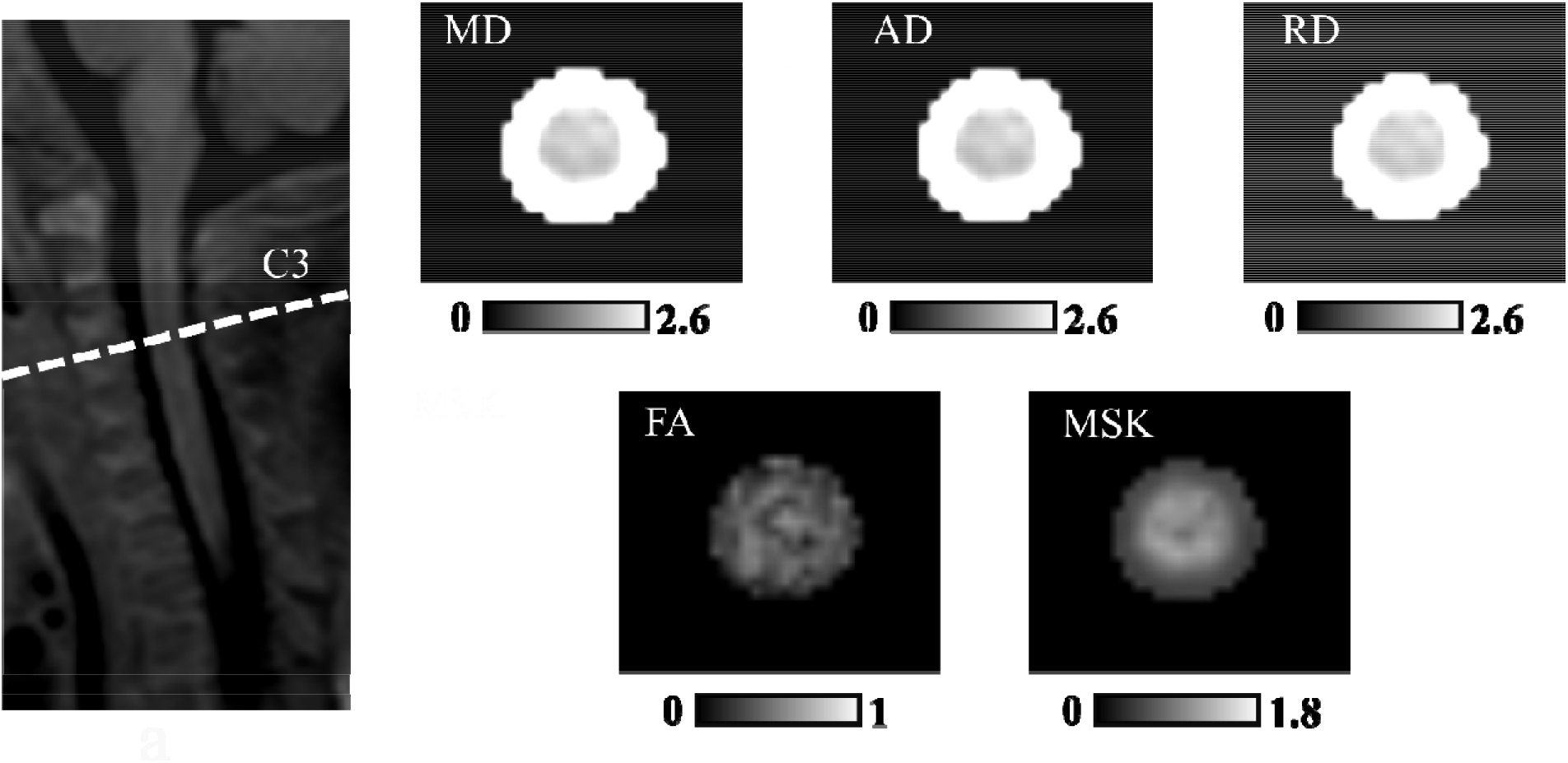
Diffusion and kurtosis maps at the mid-C3 level for one example subject: Units for MD, AD and RD are μm^2^/s, for MSK mm^2^/s, while FA is dimensionless.

**Figure 2S.**
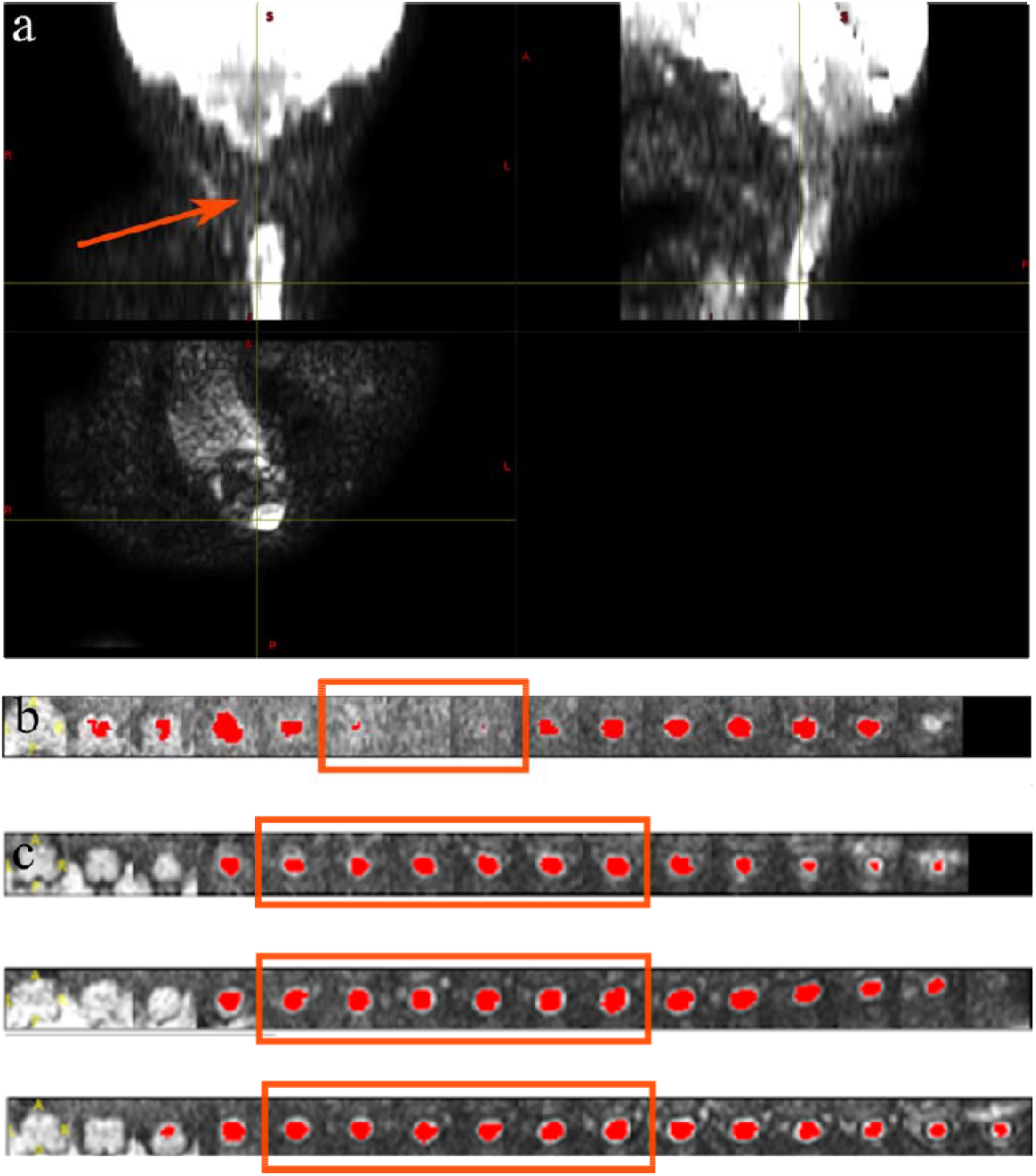
Quality control: (a) Example of excluded DKI scan and (b) relative SC segmentation show signal loss across multiple slices as the coronal plane is not overlapping with the cord (ie: lordosis); (c) QC of C1-C4 levels: axial slices under analysis correspond to the same cervical levels for all subjects as shown in three example subjects.

**Figure 3S.**
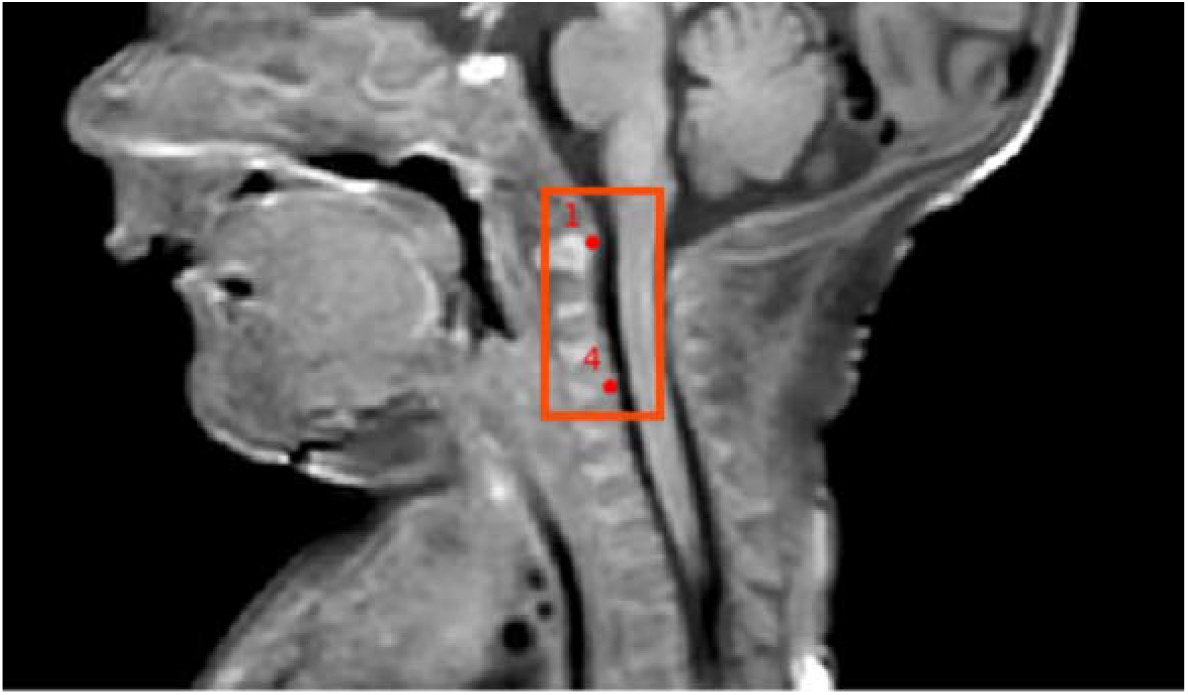
Vertebral labeling: Manual labeling of top of C1 vertebra and C3-C4 disc from graphical user interface integrated in SCT

**Figure 4S.**
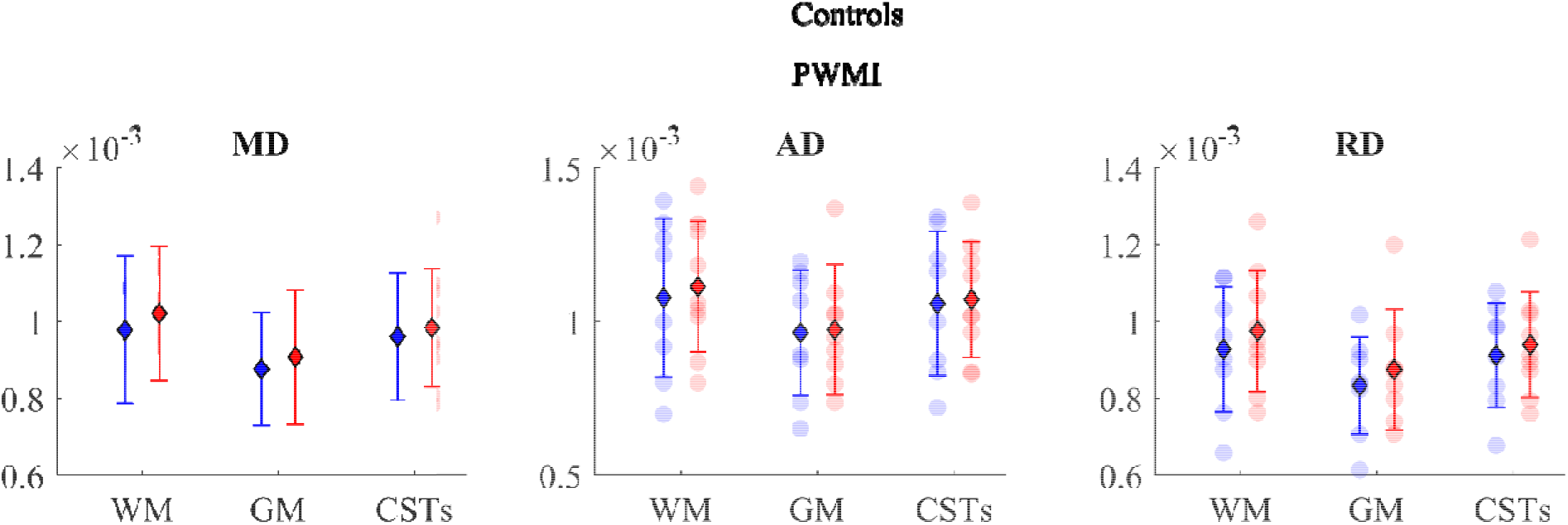
Extraction of diffusion measures within specific ROIs: Scatter plots of DTI in group subjects across aforementioned ROIs: coloured spots indicate single subject’s value for each metric; as reported in the legend, controls’ measures are in blue, whereas Periventricular White Matter Injury (PWMI) group’s in red. Units for MD, AD, RD are in mm^2^/s. Error bars displaying mean (diamond) and standard deviation (bars) are overlaid on scatter plots.

## References

1. Tournier JD. Diffusion MRI in the brain – Theory and concepts. Progress in Nuclear Magnetic Resonance Spectroscopy (2019) doi:10.1016/j.pnmrs.2019.03.001

2. Jensen JH, Helpern JA, Ramani A, Lu H, Kaczynski K. Diffusional kurtosis imaging: The quantification of non-Gaussian water diffusion by means of magnetic resonance imaging. Magnetic Resonance in Medicine (2005) doi:10.1002/mrm.20508

3. Farquharson S, Tournier JD, Calamante F, Fabinyi G, Schneider-Kolsky M, Jackson GD, Connelly A. White matter fiber tractography: Why we need to move beyond DTI. Journal of Neurosurgery (2013) doi:10.3171/2013.2.JNS121294

4. Loucao R, Nunes RG, Neto-Henriques R, Correia M, Ferreira H. Human brain tractography: A DTI vs DKI comparison analysis. in doi:10.1109/enbeng.2015.7088820

5. Mader I, Urbach H. Walk the line: From diffusion imaging to the microstructure of the brain. Clinical Neuroradiology (2013) doi:10.1007/s00062-013-0265-3

6. Cohen Y, Anaby D, Morozov D. Diffusion MRI of the spinal cord: from structural studies to pathology. NMR in Biomedicine (2017) doi:10.1002/nbm.3592

7. Wu EX, Cheung MM. MR diffusion kurtosis imaging for neural tissue characterization. NMR in Biomedicine (2010) doi:10.1002/nbm.1506

8. Taber KH, Herrick RC, Weathers SW, Kumar A, Schomer DF, Hayman LA. Pitfalls and Encountered Clinical Artifacts in Imaging of the Spine1. Radiographics□: a review publication of the Radiological Society of North America, Inc (1998)

9. Panara V, Navarra R, Mattei PA, Piccirilli E, Cotroneo AR, Papinutto N, Henry RG, Uncini A, Caulo M. Spinal cord microstructure integrating phase-sensitive inversion recovery and diffusional kurtosis imaging. Neuroradiology (2017) doi:10.1007/s00234-017-1864-5

10. Li D, Wang X. Application value of diffusional kurtosis imaging (DKI) in evaluating microstructural changes in the spinal cord of patients with early cervical spondylotic myelopathy. Clinical Neurology and Neurosurgery (2017) doi:10.1016/j.clineuro.2017.03.015

11. Bester M, Sigmund E, Tabesh A, Jaggi H, Inglese M, Mitnick R. Diffusional Kurtosis Imaging of the cervical spinal cord in multiple sclerosis patients. Proc Intl Soc Mag Reson Med (2010)

12. Bester M, Sigmund E, Tabesh A, Jaggi H, Inglese M, Mitnick R. Diffusional Kurtosis Imaging of the cervical spinal cord in multiple sclerosis patients. in

13. Raz E, Bester M, Sigmund EE, Tabesh A, Babb JS, Jaggi H, Helpern J, Mitnick RJ, Inglese M. A better characterization of spinal cord damage in multiple sclerosis: A diffusional kurtosis imaging study. in American Journal of Neuroradiology doi:10.3174/ajnr.A3512

14. Li D-W, Wang X-M. Progresses of diffusion kurtosis imaging in spinal cord injury. (2015) 31:1422–1425. doi:10.13929/j.1003-3289.2015.09.036

15. Sorantin E, Robl T, Lindbichler F, Riccabona M. MRI of the neonatal and paediatric spine and spinal canal. European Journal of Radiology (2008) doi:10.1016/j.ejrad.2008.06.032

16. Thukral BB. Problems and preferences in pediatric imaging. Indian Journal of Radiology and Imaging (2015) doi:10.4103/0971-3026.169466

17. Wilm BJ, Svensson J, Henning A, Pruessmann KP, Boesiger P, Kollias SS. Reduced field-of-view MRI using outer volume suppression for spinal cord diffusion imaging. Magnetic Resonance in Medicine (2007) doi:10.1002/mrm.21167

18. Toselli B, Tortora D, Severino M, Arnulfo G, Canessa A, Morana G, Rossi A, Fato MM. Improvement in white matter tract reconstruction with constrained spherical deconvolution and track density mapping in low angular resolution data: A pediatric study and literature review. Frontiers in Pediatrics (2017) doi:10.3389/fped.2017.00182

19. Webster JG, Descoteaux M. “High Angular Resolution Diffusion Imaging (HARDI),” in Wiley Encyclopedia of Electrical and Electronics Engineering doi:10.1002/047134608x.w8258

20. Andre JB, Bammer R. Advanced diffusion-weighted magnetic resonance imaging techniques of the human spinal cord. Topics in Magnetic Resonance Imaging (2010) doi:10.1097/RMR.0b013e31823e65a1

21. Fruehwald-Pallamar J, Szomolanyi P, Fakhrai N, Lunzer A, Weber M, Thurnher MM, Pallamar M, Trattnig S, Prayer D, Noebauer-Huhmann IM. Parallel imaging of the cervical spine at 3T: Optimized trade-off between speed and image quality. American Journal of Neuroradiology (2012) doi:10.3174/ajnr.A3101

22. Saksena S, Alizadeh M, Middleton DM, Conklin CJ, Krisa L, Flanders A, Mulcahey M, Mohamed FB, Faro SH. Characterization of spinal cord diffusion tensor imaging metrics in clinically asymptomatic pediatric subjects with incidental congenital lesions. Spinal Cord Series and Cases (2018) doi:10.1038/s41394-018-0073-8

23. Alizadeh M, Fisher J, Saksena S, Sultan Y, Conklin CJ, Middleton DM, Krisa L, Finsterbusch J, Flanders AE, Faro SH, et al. Age related diffusion and tractography changes in typically developing pediatric cervical and thoracic spinal cord. NeuroImage: Clinical (2018) doi:10.1016/j.nicl.2018.03.014

24. Alizadeh M, Fisher J, Saksena S, Sultan Y, Conklin CJ, Middleton DM, Finsterbusch J, Krisa L, Flanders AE, Faro SH, et al. Reduced Field of View Diffusion Tensor Imaging and Fiber Tractography of the Pediatric Cervical and Thoracic Spinal Cord Injury. Journal of Neurotrauma (2018) doi:10.1089/neu.2017.5174

25. Mulcahey MJ, Samdani AF, Gaughan JP, Barakat N, Faro S, Shah P, Betz RR, Mohamed FB. Diagnostic accuracy of diffusion tensor imaging for pediatric cervical spinal cord injury. Spinal Cord (2013) doi:10.1038/sc.2013.36

26. Saksena S, Middleton DM, Krisa L, Shah P, Faro SH, Sinko R, Gaughan J, Finsterbusch J, Mulcahey MJ, Mohamed FB. Diffusion tensor imaging of the normal cervical and thoracic pediatric spinal cord. American Journal of Neuroradiology (2016) doi:10.3174/ajnr.A4883

27. Mohamed FB, Hunter LN, Barakat N, Liu CSJ, Sair H, Samdani AF, Betz RR, Faro SH, Gaughan J, Mulcahey MJ. Diffusion tensor imaging of the pediatric spinal cord at 1.5T: Preliminary results. American Journal of Neuroradiology (2011) doi:10.3174/ajnr.A2334

28. Antherieu P, Levy R, de Saint Denis T, Lohkamp L, Paternoster G, di Rocco F, Boddaert N, Zerah M. Diffusion tensor imaging (DTI) and Tractography of the spinal cord in pediatric population with spinal lipomas: preliminary study. Child’s Nervous System (2019) doi:10.1007/s00381-018-3935-2

29. Reynolds BB, By S, Weinberg QR, Witt AA, Newton AT, Feiler HR, Ramkorun B, Clayton DB, Couture P, Martus JE, et al. Quantification of DTI in the pediatric spinal cord: Application to clinical evaluation in a healthy patient population. American Journal of Neuroradiology (2019) doi:10.3174/ajnr.A6104

30. Singhi S, Tekes A, Thurnher M, Gilson WD, Izbudak I, Thompson CB, Huisman TAGM. Diffusion tensor imaging of the maturing paediatric cervical spinal cord: From the neonate to the young adult. Journal of Neuroradiology (2012) doi:10.1016/j.neurad.2011.05.002

31. Saksena S, Mohamed FB, Middleton DM, Krisa L, Alizadeh M, Shahrampour S, Conklin CJ, Flanders A, Finsterbusch J, Mulcahey MJ, et al. Diffusion Tensor Imaging Assessment of Regional White Matter Changes in the Cervical and Thoracic Spinal Cord in Pediatric Subjects. Journal of Neurotrauma (2018) 36:853–861. doi:10.1089/neu.2018.5826

32. Conklin CJ, Middleton DM, Alizadeh M, Finsterbusch J, Raunig DL, Faro SH, Shah P, Krisa L, Sinko R, Delalic JZ, et al. Spatially selective 2D RF inner field of view (iFOV) diffusion kurtosis imaging (DKI) of the pediatric spinal cord. NeuroImage: Clinical (2016) doi:10.1016/j.nicl.2016.01.009

33. Singh G, True AJ, Lui CC, Prasanna P, Orleans G, Partyka L, Phatak TD. Normal anterior-posterior diameters of the spinal cord and spinal canal in healthy term newborns on sonography. Pediatric Radiology (2020) doi:10.1007/s00247-020-04879-8

34. Kaplan KM, Spivak JM, Bendo JA. Embryology of the spine and associated congenital abnormalities. Spine Journal (2005) doi:10.1016/j.spinee.2004.10.044

35. Oskouian RJ, Sansur CA, Shaffrey CI. Congenital Abnormalities of the Thoracic and Lumbar Spine. Neurosurgery Clinics of North America (2007) doi:10.1016/j.nec.2007.04.004

36. Rufener S, Ibrahim M, Parmar HA. Imaging of Congenital Spine and Spinal Cord Malformations. Neuroimaging Clinics of North America (2011) doi:10.1016/j.nic.2011.05.011

37. S Basu P, Elsebaie H, Noordeen M. Congenital spinal deformity: A comprehensive assessment at presentation. Spine (2002) doi:10.1097/00007632-200210150-00014

38. Yang RK, Roth CG, Ward RJ, deJesus JO, Mitchell DG. Optimizing abdominal MR imaging: Approaches to common problems. Radiographics (2010) doi:10.1148/rg.301095076

39. Tournier JD, Smith R, Raffelt D, Tabbara R, Dhollander T, Pietsch M, Christiaens D, Jeurissen B, Yeh CH, Connelly A. MRtrix3: A fast, flexible and open software framework for medical image processing and visualisation. NeuroImage (2019) doi:10.1016/j.neuroimage.2019.116137

40. de Leener B, Lévy S, Dupont SM, Fonov VS, Stikov N, Louis Collins D, Callot V, Cohen-Adad J. SCT: Spinal Cord Toolbox, an open-source software for processing spinal cord MRI data. NeuroImage (2017) doi:10.1016/j.neuroimage.2016.10.009

41. Garyfallidis E, Brett M, Amirbekian B, Rokem A, van der Walt S, Descoteaux M, Nimmo-Smith I. Dipy, a library for the analysis of diffusion MRI data. Frontiers in Neuroinformatics (2014) doi:10.3389/fninf.2014.00008

42. Battiston M, Grussu F, Ianus A, Schneider T, Prados F, Fairney J, Ourselin S, Alexander DC, Cercignani M, Gandini Wheeler-Kingshott CAM, et al. An optimized framework for quantitative magnetization transfer imaging of the cervical spinal cord in vivo. Magnetic Resonance in Medicine (2018) doi:10.1002/mrm.26909

43. Brooks JCW, Büchel C, Winkler AM, Andersson JL, Tracey I. Investigating resting-state functional connectivity in the cervical spinal cord at 3 T. NeuroImage (2017) doi:10.1016/j.neuroimage.2016.12.072

44. Duval T, Lévy S, Stikov N, Cohen-Adad J, Stikov N, Campbell J, Mezer A, Witzel T, Keil B, Smith V, et al. g-Ratio weighted imaging of the human spinal cord in vivo. NeuroImage (2017) doi:10.1016/j.neuroimage.2016.09.018

45. Eippert F, Kong Y, Jenkinson M, Tracey I, Brooks JCW. Denoising spinal cord fMRI data: Approaches to acquisition and analysis. NeuroImage (2017) doi:10.1016/j.neuroimage.2016.09.065

46. Kong Y, Eippert F, Beckmann CF, Andersson J, Finsterbusch J, Büchel C, Tracey I, Brooks JCW. Intrinsically organized resting state networks in the human spinal cord. Proceedings of the National Academy of Sciences of the United States of America (2014) doi:10.1073/pnas.1414293111

47. Samson RS, Lévy S, Schneider T, Smith AK, Smith SA, Cohen-Adad J, Wheeler-Kingshott CAMG. ZOOM or Non-ZOOM? Assessing spinal cord diffusion tensor imaging protocols for multi-centre studies. PLoS ONE (2016) doi:10.1371/journal.pone.0155557

48. Massire A, Taso M, Besson P, Guye M, Ranjeva JP, Callot V. High-resolution multi-parametric quantitative magnetic resonance imaging of the human cervical spinal cord at 7T. NeuroImage (2016) doi:10.1016/j.neuroimage.2016.08.055

49. Taso M, Girard OM, Duhamel G, le Troter A, Feiweier T, Guye M, Ranjeva JP, Callot V. Tract-specific and age-related variations of the spinal cord microstructure: A multi-parametric MRI study using diffusion tensor imaging (DTI) and inhomogeneous magnetization transfer (ihMT). NMR in Biomedicine (2016) doi:10.1002/nbm.3530

50. Vahdat S, Lungu O, Cohen-Adad J, Marchand-Pauvert V, Benali H, Doyon J. Simultaneous brain–cervical cord fMRI reveals intrinsic spinal cord plasticity during motor sequence learning. PLoS Biology (2015) doi:10.1371/journal.pbio.1002186

51. Weber KA, Sentis AI, Bernadel-Huey ON, Chen Y, Wang X, Parrish TB, Mackey S. Thermal Stimulation Alters Cervical Spinal Cord Functional Connectivity in Humans. Neuroscience (2018) doi:10.1016/j.neuroscience.2017.10.035

52. Weber KA, Chen Y, Wang X, Kahnt T, Parrish TB. Lateralization of cervical spinal cord activity during an isometric upper extremity motor task with functional magnetic resonance imaging. NeuroImage (2016) doi:10.1016/j.neuroimage.2015.10.014

53. Ljungberg E, Vavasour I, Tam R, Yoo Y, Rauscher A, Li DKB, Traboulsee A, MacKay A, Kolind S. Rapid myelin water imaging in human cervical spinal cord. Magnetic Resonance in Medicine (2017) doi:10.1002/mrm.26551

54. Castellano A, Papinutto N, Cadioli M, Brugnara G, Iadanza A, Scigliuolo G, Pareyson D, Uziel G, Köhler W, Aubourg P, et al. Quantitative MRI of the spinal cord and brain in adrenomyeloneuropathy: In vivo assessment of structural changes. Brain (2016) doi:10.1093/brain/aww068

55. Grabher P, Mohammadi S, Trachsler A, Friedl S, David G, Sutter R, Weiskopf N, Thompson AJ, Curt A, Freund P. Voxel-based analysis of grey and white matter degeneration in cervical spondylotic myelopathy. Scientific Reports (2016) doi:10.1038/srep24636

56. Hori M, Hagiwara A, Fukunaga I, Ueda R, Kamiya K, Suzuki Y, Liu W, Murata K, Takamura T, Hamasaki N, et al. Application of Quantitative Microstructural MR Imaging with Atlas-based Analysis for the Spinal Cord in Cervical Spondylotic Myelopathy. Scientific Reports (2018) doi:10.1038/s41598-018-23527-8

57. Huber E, David G, Thompson AJ, Weiskopf N, Mohammadi S, Freund P. Dorsal and ventral horn atrophy is associated with clinical outcome after spinal cord injury. Neurology (2018) doi:10.1212/WNL.0000000000005361

58. McCoy DB, Talbott JF, Wilson M, Mamlouk MD, Cohen-Adad J, Wilson M, Narvid J. MRI atlas-based measurement of spinal cord injury predicts outcome in acute flaccid myelitis. American Journal of Neuroradiology (2017) doi:10.3174/ajnr.A5044

59. Martin AR, de Leener B, Cohen-Adad J, Cadotte DW, Kalsi-Ryan S, Lange SF, Tetreault L, Nouri A, Crawley A, Mikulis DJ, et al. A novel MRI biomarker of spinal cord white matter injury: T2∗-weighted white matter to gray matter signal intensity ratio. American Journal of Neuroradiology (2017) doi:10.3174/ajnr.A5162

60. Smith AC, Weber KA, O’Dell DR, Parrish TB, Wasielewski M, Elliott JM. Lateral Corticospinal Tract Damage Correlates With Motor Output in Incomplete Spinal Cord Injury. Archives of Physical Medicine and Rehabilitation (2018) doi:10.1016/j.apmr.2017.10.002

61. Talbott JF, Narvid J, Chazen JL, Chin CT, Shah V. An Imaging-Based Approach to Spinal Cord Infection. Seminars in Ultrasound, CT and MRI (2016) doi:10.1053/j.sult.2016.05.006

62. Ventura RE, Kister I, Chung S, Babb JS, Shepherd TM. Cervical spinal cord atrophy in nmosd without a history of myelitis or MRI-visible lesions. Neurology: Neuroimmunology and NeuroInflammation (2016) doi:10.1212/NXI.0000000000000224

63. Yiannakas MC, Mustafa AM, de Leener B, Kearney H, Tur C, Altmann DR, de Angelis F, Plantone D, Ciccarelli O, Miller DH, et al. Fully automated segmentation of the cervical cord from T1-weighted MRI using PropSeg: Application to multiple sclerosis. NeuroImage: Clinical (2016) doi:10.1016/j.nicl.2015.11.001

64. Barth M, Breuer F, Koopmans PJ, Norris DG, Poser BA. Imaging Methodology - Review Simultaneous Multislice (SMS) Imaging Techniques. Magnetic Resonance in Medicine (2016)

65. Fadnavis S, Batson J, Garyfallidis E. Patch2Self: Denoising Diffusion MRI with Self-Supervised Learning. (2020) 1–11. Available at: http://arxiv.org/abs/2011.01355

66. Schilling K, Fadnavis S, Visagie M, Garyfallidis E, Landman B, Smith S, O’grady K. Patch2Self denoising of diffusion MRI in the cervical spinal cord improves repeatability and feature conspicuity. in

67. Xu J, Shimony JS, Klawiter EC, Snyder AZ, Trinkaus K, Naismith RT, Benzinger TLS, Cross AH, Song SK. Improved in vivo diffusion tensor imaging of human cervical spinal cord. NeuroImage (2013) doi:10.1016/j.neuroimage.2012.11.014

68. de Leener B, Kadoury S, Cohen-Adad J. Robust, accurate and fast automatic segmentation of the spinal cord. NeuroImage (2014) doi:10.1016/j.neuroimage.2014.04.051

69. Gros C, de Leener B, Badji A, Maranzano J, Eden D, Dupont SM, Talbott J, Zhuoquiong R, Liu Y, Granberg T, et al. Automatic segmentation of the spinal cord and intramedullary multiple sclerosis lesions with convolutional neural networks. NeuroImage (2019) doi:10.1016/j.neuroimage.2018.09.081

70. Ullmann E, Pelletier Paquette JF, Thong WE, Cohen-Adad J. Automatic Labeling of Vertebral Levels Using a Robust Template-Based Approach. International Journal of Biomedical Imaging (2014) doi:10.1155/2014/719520

71. de Leener B, Fonov VS, Collins DL, Callot V, Stikov N, Cohen-Adad J. PAM50: Unbiased multimodal template of the brainstem and spinal cord aligned with the ICBM152 space. NeuroImage (2018) doi:10.1016/j.neuroimage.2017.10.041

72. de Leener B, Mangeat G, Dupont S, Martin AR, Callot V, Stikov N, Fehlings MG, Cohen-Adad J. Topologically preserving straightening of spinal cord MRI. Journal of Magnetic Resonance Imaging (2017) doi:10.1002/jmri.25622

73. Tustison NJ, Avants BB. Explicit B-spline regularization in diffeomorphic image registration. Frontiers in Neuroinformatics (2013) doi:10.3389/fninf.2013.00039

74. Henriques RN, Correia MM, Marrale M, Huber E, Kruper J, Koudoro S, Yeatman JD, Garyfallidis E, Rokem A. Diffusional Kurtosis Imaging in the Diffusion Imaging in Python Project. Frontiers in Human Neuroscience (2021) doi:10.3389/fnhum.2021.675433

75. Lévy S, Benhamou M, Naaman C, Rainville P, Callot V, Cohen-Adad J. White matter atlas of the human spinal cord with estimation of partial volume effect. NeuroImage (2015) doi:10.1016/j.neuroimage.2015.06.040

76. Tax CMW, Otte WM, Viergever MA, Dijkhuizen RM, Leemans A. REKINDLE: Robust Extraction of Kurtosis INDices with Linear Estimation. Magnetic Resonance in Medicine (2015) doi:10.1002/mrm.25165

77. Neto Henriques R. Diffusion kurtosis imaging of the healthy human brain. (2012) 134. Available at: http://repositorio.ul.pt/handle/10451/8511

78. Henriques RN, Jespersen SN, Jones DK, Veraart J. Toward more robust and reproducible diffusion kurtosis imaging. Magnetic Resonance in Medicine (2021) doi:10.1002/mrm.28730

79. Henriques RN. Advanced Methods for Diffusion MRI Data Analysis and their Application to the Healthy Ageing Brain. (2017)

80. Neto Henriques R, Correia MM, Nunes RG, Ferreira HA. Exploring the 3D geometry of the diffusion kurtosis tensor-Impact on the development of robust tractography procedures and novel biomarkers. NeuroImage (2015) doi:10.1016/j.neuroimage.2015.02.004

81. Henriques RN, Jespersen SN, Shemesh N. Microscopic anisotropy misestimation in spherical-mean single diffusion encoding MRI. Magnetic Resonance in Medicine (2019) doi:10.1002/mrm.27606

82. Panara V, Navarra R, Mattei PA, Piccirilli E, Bartoletti V, Uncini A, Caulo M. Correlations between cervical spinal cord magnetic resonance diffusion tensor and diffusion kurtosis imaging metrics and motor performance in patients with chronic ischemic brain lesions of the corticospinal tract. Neuroradiology (2019) doi:10.1007/s00234-018-2139-5

83. Volpe JJ, Inder TE, Darras BT, de Vries LS, du Plessis AJ, Neil JJ, Perlman J. Volpe’s neurology of the newborn. (2017). doi:10.1016/c2010-0-68825-0

84. Veraart J, Novikov DS, Christiaens D, Ades-aron B, Sijbers J, Fieremans E. Denoising of diffusion MRI using random matrix theory. NeuroImage (2016) doi:10.1016/j.neuroimage.2016.08.016

85. Hastie T, Tibshirani R, Friedman J. The Elements of Statistical Learning Data Mining, Inference, and Prediction (12th printing). (2017).

86. Pierpaoli C, Basser PJ. Toward a quantitative assessment of diffusion anisotropy. Magnetic Resonance in Medicine (1996) doi:10.1002/mrm.1910360612

87. Pierpaoli C, Jezzard P, Basser PJ, Barnett A, di Chiro G. Diffusion tensor MR imaging of the human brain. Radiology (1996) doi:10.1148/radiology.201.3.8939209

88. Parodi A, Malova M, Cardiello V, Raffa S, Re M, Calevo MG, Severino M, Tortora D, Morana G, Rossi A, et al. Punctate white matter lesions of preterm infants: Risk factor analysis. European Journal of Paediatric Neurology (2019) doi:10.1016/j.ejpn.2019.06.003

89. Cheong JLY, Thompson DK, Wang HX, Hunt RW, Anderson PJ, Inder TE, Doyle LW. Abnormal white matter signal on MR imaging is related to abnormal tissue microstructure. American Journal of Neuroradiology (2009) doi:10.3174/ajnr.A1399

90. Staudt M, Pavlova M, Böhm S, Grodd W, Krägeloh-Mann I. Pyramidal tract damage correlates with motor dysfunction in bilateral periventricular leukomalacia (PVL). Neuropediatrics (2003) doi:10.1055/s-2003-42206

91. Bassi L, Chew A, Merchant N, Ball G, Ramenghi L, Boardman J, Allsop JM, Doria V, Arichi T, Mosca F, et al. Diffusion tensor imaging in preterm infants with punctate white matter lesions. Pediatric Research (2011) doi:10.1203/PDR.0b013e3182182836

92. Tusor N, Benders MJ, Counsell SJ, Nongena P, Ederies MA, Falconer S, Chew A, Gonzalez-Cinca N, Hajnal J v., Gangadharan S, et al. Punctate White Matter Lesions Associated with Altered Brain Development and Adverse Motor Outcome in Preterm Infants. Scientific Reports (2017) doi:10.1038/s41598-017-13753-x

93. Paydar A, Fieremans E, Nwankwo JI, Lazar M, Sheth HD, Adisetiyo V, Helpern JA, Jensen JH, Milla SS. Diffusional kurtosis imaging of the developing brain. American Journal of Neuroradiology (2014) doi:10.3174/ajnr.A3764

94. Hansen B. An Introduction to Kurtosis Fractional Anisotropy. AJNR American journal of neuroradiology (2019) 40:1638–1641. doi:10.3174/ajnr.A6235

95. Zhang H, Schneider T, Wheeler-Kingshott CA, Alexander DC. NODDI: Practical in vivo neurite orientation dispersion and density imaging of the human brain. NeuroImage (2012) doi:10.1016/j.neuroimage.2012.03.072

